# Increased interregional virus exchange and nucleotide diversity outline the expansion of the chikungunya virus ECSA lineage in Brazil

**DOI:** 10.1101/2023.03.28.23287733

**Authors:** Joilson Xavier, Luiz Alcantara, Vagner Fonseca, Mauricio Lima, Emerson Castro, Hegger Fritsch, Carla Oliveira, Natalia Guimarães, Talita Adelino, Mariane Evaristo, Evandra S. Rodrigues, Elaine Vieira Santos, Debora de La-Roque, Laise de Moraes, Stephane Tosta, Adelino Neto, Alexander Rosewell, Ana Flavia Mendonça, Anderson Leite, Andreza Vasconcelos, Arabela L. Silva de Mello, Bergson Vasconcelos, Camila A. Montalbano, Camila Zanluca, Carla Freitas, Carlos F. C. de Albuquerque, Claudia Nunes Duarte dos Santos, Cleiton S. Santos, Cliomar Alves dos Santos, Crhistinne C. Maymone Gonçalves, Dalane Teixeira, Daniel F. L. Neto, Diego Cabral, Elaine C. de Oliveira, Ethel L. Noia Maciel, Felicidade Mota Pereira, Felipe Iani, Fernanda P. de Carvalho, Gabriela Andrade, Gabriela Bezerra, Gislene G. de Castro Lichs, Glauco Carvalho Pereira, Haline Barroso, Helena Cristina Ferreira Franz, Hivylla Ferreira, Iago Gomes, Irina N. Riediger, Isabela Rodrigues, Isadora C. de Siqueira, Jacilane Silva, Jairo Mendez Rico, Jaqueline Lima, Jayra Abrantes, Jean Phellipe M. do Nascimento, Judith N. Wasserheit, Julia Pastor, Jurandy J. F. de Magalhães, Kleber Giovanni Luz, Lidio G. Lima Neto, Livia C. V. Frutuoso, Luana Barbosa da Silva, Ludmila Sena, Luis Arthur F. de Sousa, Luiz Augusto Pereira, Luiz Demarchi, Magaly C. B. Câmara, Marcela G. Astete, Maria Almiron, Maricelia Lima, Marina C. S. Umaki Zardin, Mayra M. Presibella, Melissa B. Falcão, Michael Gale, Naishe Freire, Nelson Marques, Noely F. O. de Moura, Pedro E. Almeida Da Silva, Peter Rabinowitz, Rivaldo V. da Cunha, Karen S. Trinta, Rodrigo F. do Carmo Said, Rodrigo Kato, Rodrigo Stabeli, Ronaldo de Jesus, Roselene Hans Santos, Simone K. Haddad, Svetoslav N. Slavov, Tamires Andrade, Themis Rocha, Thiago Carneiro, Vanessa Nardy, Vinicius da Silva, Walterlene G. Carvalho, Wesley C. Van Voorhis, Wildo N. Araujo, Ana M.B. de Filippis, Marta Giovanetti

**Affiliations:** Instituto Rene Rachou, Fundação Oswaldo Cruz, Minas Gerais, Brazil; Instituto de Ciências Biológicas, Universidade Federal de Minas Gerais, Brazil; Organização Pan-Americana da Saúde, Organização Mundial da Saúde, Brazil; Laboratório Central de Saúde Pública de Minas Gerais, Fundação Ezequiel Dias, Brazil; Instituto Oswaldo Cruz, Fundação Oswaldo Cruz, Rio de Janeiro, Brazil; Fundação Hemocentro de Ribeirão Preto, Brazil; Instituto Gonçalo Moniz, Fundação Oswaldo Cruz, Bahia, Brazil; Laboratório Central de Saúde Pública do Piaui, Brazil; Laboratório Central de Saúde Pública de Goias, Brazil; Laboratório Central de Saúde Pública de Alagoas, Brazil; Laboratório Central de Saúde Pública de Pernambuco, Brazil; Laboratório Central de Saúde Pública da Bahia, Brazil; Laboratório Central de Saúde Pública da Paraíba, Brazil; Universidade Federal de Mato Grosso do Sul; Instituto Carlos Chagas, Fundação Oswaldo Cruz, Paraná, Brazil; Coordenação Geral dos Laboratórios de Saúde Pública, Ministério da Saúde, Brazil; Laboratório Central de Saúde Pública de Sergipe, Brazil; Secretaria de Saúde do Estado do Mato Grosso do Sul, Brazil; Laboratório Central de Saúde Pública do Mato Grosso, Brazil; Secretaria de Vigilância em Saúde e Ambiente, Ministério da Saúde, Brazil; Laboratório Central de Saúde Pública do Mato Grosso do Sul, Brazil; Laboratório Central de Saúde Pública do Maranhão, Brazil; Laboratório Central de Saúde Pública do Rio Grande do Norte, Brazil; Laboratório Central de Saúde Pública do Paraná, Brazil; Pan American Health Organization, Washington, USA; Department of Global Health and Medicine, University of Washington, USA; Universidade de Pernambuco Campus Serra Talhada; Universidade Federal do Rio Grande do Norte; Coordenação Geral das Arboviroses, Ministério da Saúde, Brazil; Universidade Estadual de Feira de Santana, Brazil; Secretaria de Saúde de Feira de Santana, Feira de Santana, Bahia, Brazil; Department of Immunology, University of Washington, USA; Department of Environmental and Occupational Health Sciences, University of Washington, USA; Fundação Oswaldo Cruz, Instituto de Tecnologia em Imunobiológicos, Brazil; Center for Research Development, CDC, Butantan Institute, Brazil; Department of Medicine, University of Washington, USA; Sciences and Technologies for Sustainable Development and One Health, University of Campus Bio-Medico, Italy

**Author notes:** Correspondence, &. These authors contributed equally (Joilson Xavier, Luiz Alcantara, Vagner Fonseca).

**Keywords:** chikungunya virus, East-Central-South-African lineage, phylogenetics, nanopore sequencing, genomic surveillance, genetic diversity

## Abstract

The emergence and reemergence of mosquito-borne diseases in Brazil such as Yellow Fever, Zika, Chikungunya, and Dengue have had serious impacts on public health. Concerns have been raised due to the rapid dissemination of the chikungunya virus (CHIKV) across the country since its first detection in 2014 in Northeast Brazil. Faced with this scenario, on-site training activities in genomic surveillance carried out in partnership with the National Network of Public Health Laboratories have led to the generation of 422 CHIKV genomes from 12 Brazilian states over the past two years (2021-2022), a period that has seen more than 312 thousand chikungunya fever cases reported in the country. These new genomes increased the amount of available data and allowed a more comprehensive characterization of the dispersion dynamics of the CHIKV East-Central-South-African (ECSA) lineage in Brazil. Tree branching patterns revealed the emergence and expansion of two distinct subclades. Phylogeographic analysis indicated that the northeast region has been the leading hub of virus spread towards other regions. Increased frequency of C>T transitions among the new genomes suggested that host restriction factors from the immune system such as ADAR and AID/APOBEC deaminases might be driving CHIKV ECSA lineage genetic diversity in Brazil.

## Introduction

Mosquito-borne viral diseases have impacted the lives of millions of people across several populations in the tropics and subtropics^1,2^. This scenario prompted the World Health Organization (WHO) to issue a guideline in 2017 with strategies for global vector control response aiming to reduce the burden and threat of vector-borne diseases, such as Dengue, Zika and Chikungunya fever, by 2030^3^. Five years later, considering the public health implications brought by the coronavirus disease (COVID-19) pandemic, the WHO outlined ten proposals to strengthen health emergency preparedness, response, and resilience^4^. One of the proposals calls for the development and establishment of a collaborative surveillance system with improved laboratory capacity for pathogen and genomic surveillance that would guide the public health response.

Several genomic surveillance initiatives have been carried out in the last few years to build knowledge regarding the genetic diversity and transmission dynamics of arboviruses in Brazil^5–11^. Genomic sequencing has revealed that the first case of chikungunya fever (CHIKF) reported in Brazil was an infection by the Asian lineage introduced in a northern state in 2014, while another case reported seven days later in a north-eastern state represented the first known introduction of the East-Central-South-African (ECSA) lineage in the country^12^. The establishment of the chikungunya virus (CHIKV) in the Brazilian territory was followed by several outbreaks reported across the country, accounting for more than 200 thousand confirmed cases in only the last two years^13^. Viral genomic data from Brazilian cases has revealed that the ECSA lineage is widespread throughout the country and has been linked to fatal cases observed in both risk and non-risk groups (young adults and no commodities)^14,15^.

Viral genomic surveillance activities have been driven by the rapid development of DNA sequencing technology and bioinformatics tools for genomic data analysis^16^. Such tools have allowed the characterization of the genome and dispersal patterns of emerging and reemerging pathogens^6,9,17^. The use of such tools during the COVID-19 pandemic allowed, for example, the rapid identification of emerging mutations likely associated with increased transmissibility and immune escape^18^.

Despite technological advances and the high number of CHIKF cases reported in recent years in Brazil, the amount of genomic data available in public databases has consisted of genomes from localized outbreaks that could be limited in terms of representativeness across different states and outbreak events. CHIKF can cause long last effects such as debilitating arthritis and arthralgia, and there are no effective treatments available^19^. Vaccine candidates in development have reached phase 2 and 3 clinical trials using attenuated virus derived from the Indian Ocean lineage (IOL) or virus-like particle containing recombinant structural proteins derived from a Senegalese viral strain^20,21^. Since available vaccine candidates are based on non-Brazilian variants, increasing the availability of genomic data to characterize the genetic pool of the viral population circulating in Brazil might facilitate the future development of an efficient and more representative CHIKV vaccine.

Recurring outbreaks demonstrate that CHIKV is currently endemic in Brazil. The existence of abundant vectors, together with adequate climatic conditions for vector survival in areas of high population density, create conditions that can modify the adaptive landscape, allowing the continued expansion and evolutionary adaptation of CHIKV^22–24^. Faced with a scenario of limited availability of genetic information on potential strains causing a rapid increase in the number of CHIKF cases over the past two years in Brazil, we carried out on-site training activities in genomic surveillance in 12 Brazilian states covering four geographic regions to increase the number of available viral genomic sequences. This has allowed comprehensive monitoring of the expansion of the ECSA lineage and its variants circulating in different states, in addition to the characterization of the most up-to-date structured phylogeny of the CHIKV ECSA lineage in the country.

## Results

### East, Central, and South African (ECSA) lineage monitoring through countrywide genomic surveillance

Nanopore sequencing was performed on selected CHIKV-positive samples provided by state public health laboratories from 12 states across 4 geographic regions (Northeast, Midwest, Southeast, and South) of Brazil (**Figure 1A**) during the years 2021-2022, which saw a significant increase in the number of CHIKF cases reported across different Brazilian regions, with a peak incidence rate of more than 20 cases per 100,000 population, and a total of more than 312,000 reported cases nationally in this two-year period (**Figure 1B**). Due to the portability and easy setup of the nanopore sequencing protocol, which allows data generation in less than 24h, the collaborative work with the public health laboratories was able to not only generate genomic data but also promote on-site genomic surveillance training activities for the local laboratory staff. This approach combining wet lab and basic data analysis training allowed local teams to understand how genomic data can be linked to demographic information in order to produce comprehensive and relevant inferences regarding the epidemiology and evolution of CHIKV circulating in Brazil.

**Figure 1.**
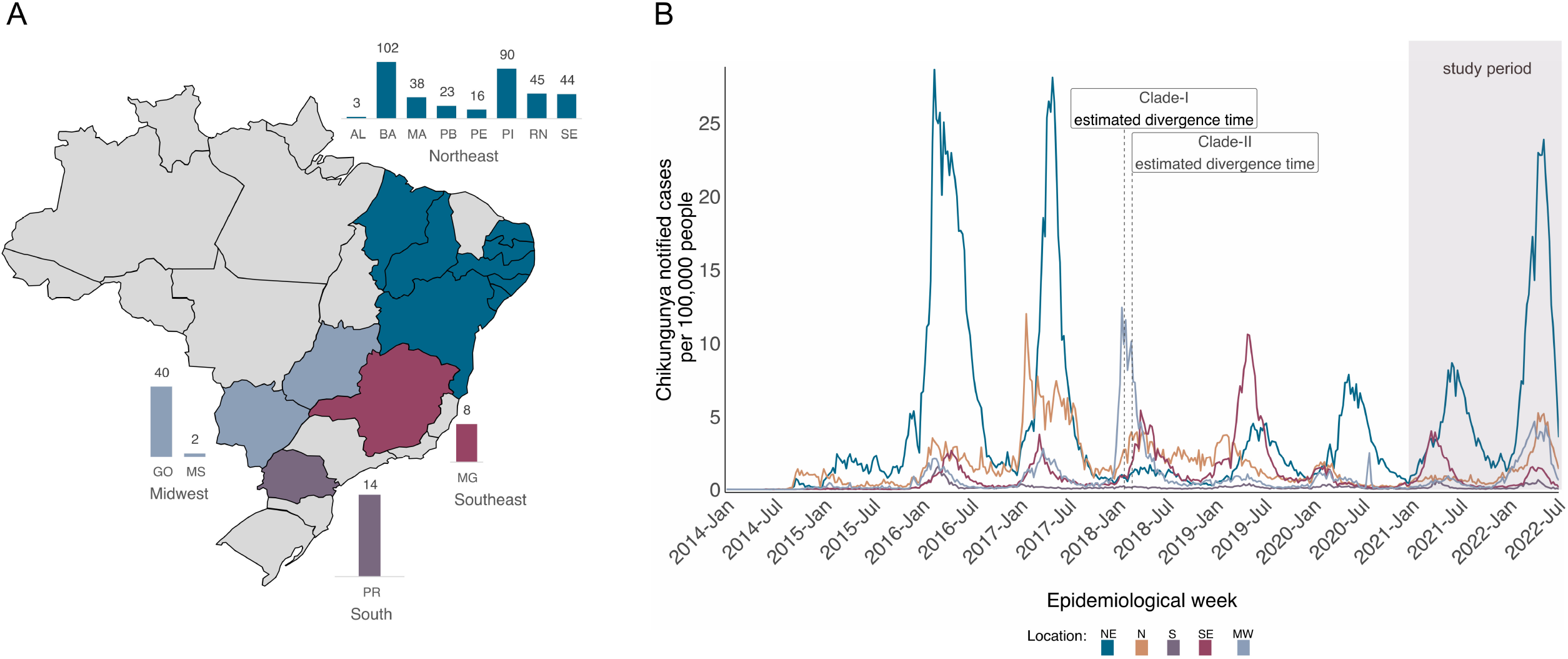
Spatiotemporal distribution of Chikungunya fever (CHIKF) in Brazil. A) Map of Brazil displays states (colored) where samples were collected and sequenced. Bar plots indicate the number of isolates obtained from each state. States abbreviations: AL=Alagoas, BA=Bahia, MA=Maranhão, PB=Paraíba, PE=Pernambuco, PI=Piauí, RN=Rio Grande do Norte, SE=Sergipe, MG=Minas Gerais, PR=Paraná, GO=Goiás, MS=Mato Grosso do Sul. B) Time series of monthly reported CHIKF cases normalized per 100□K individuals in five Brazilian macroregions over 2014-2022 (until epidemiological week 28). Epidemic curves are colored according to geographical macroregion: N□=□North, NE□=□Northeast, MW□=□Midwest, SE□=□Southeast, S□=□South. The shaded rectangle indicates the period in which samples were collected for this study.

A total of 425 CHIKV-positive samples were subjected to nanopore whole-genome sequencing, with 84.94% (n=361) of these samples originating in the Northeast region, consisting largely of samples collected from the state of Bahia (n=102) (**Figure 1A** and **Table 1**). These samples presented a mean RT-qPCR cycle threshold value of 24.04 (ranging from 11 to 35.90) (**Table 2**). Patients’ mean age upon sample collection was similar for both females and males (39 years of age), with 57.88% (n=246) of the participants identified as female (**Table 2**). The clinical status of patients at the time of sample collection, and travel history data were not available for these samples.

**Table 1:**
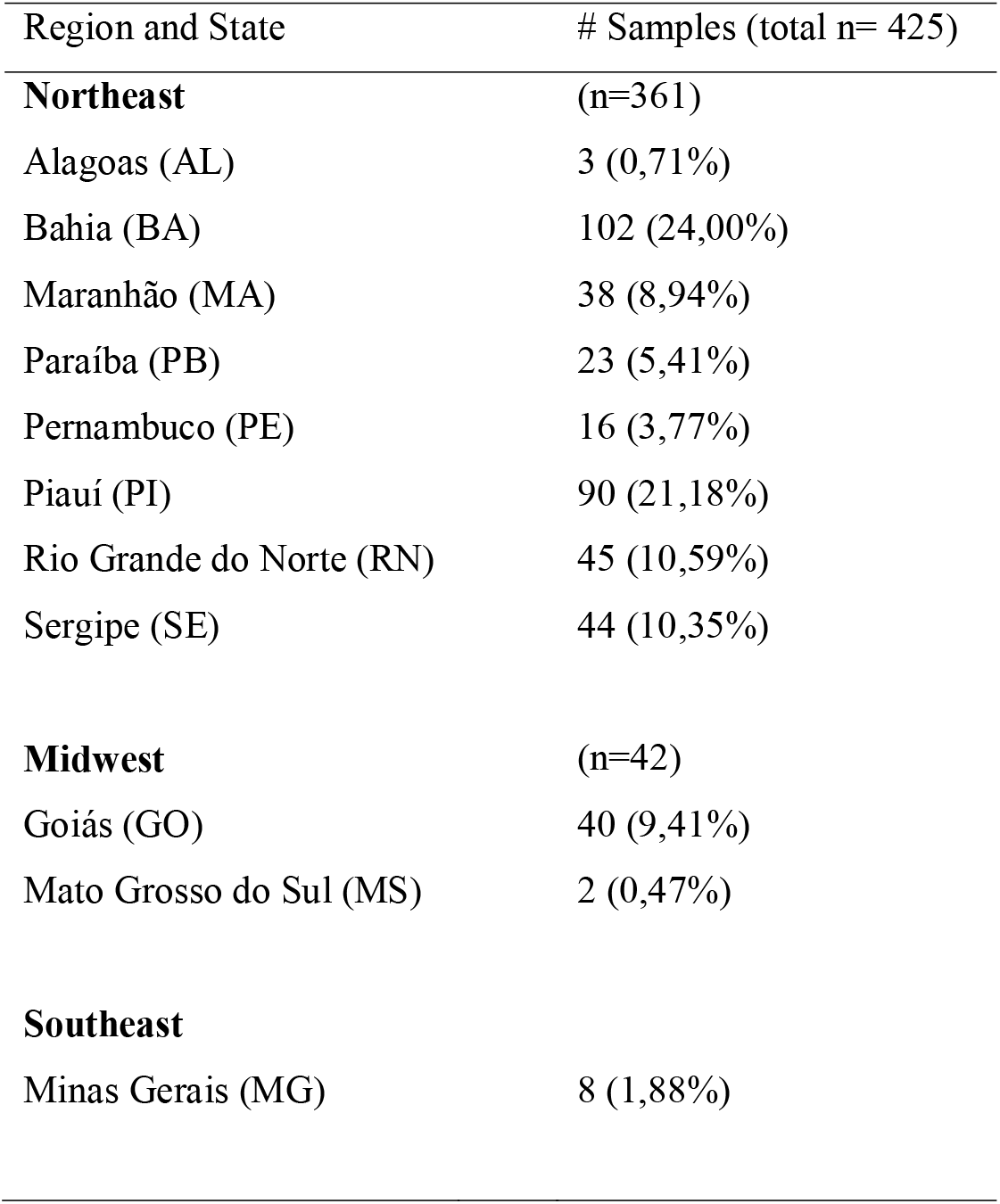
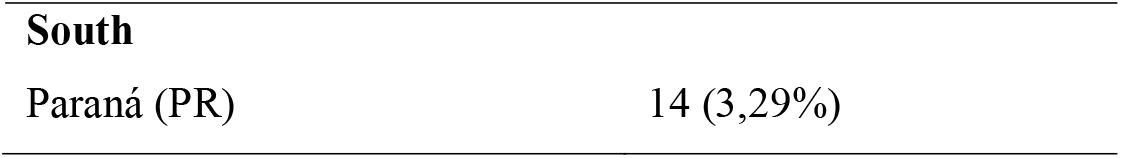
Number of Chikungunya virus-positive samples sequenced during the genomic surveillance in Brazil, 2021-2022, by geographical origin.

**Table 2:**
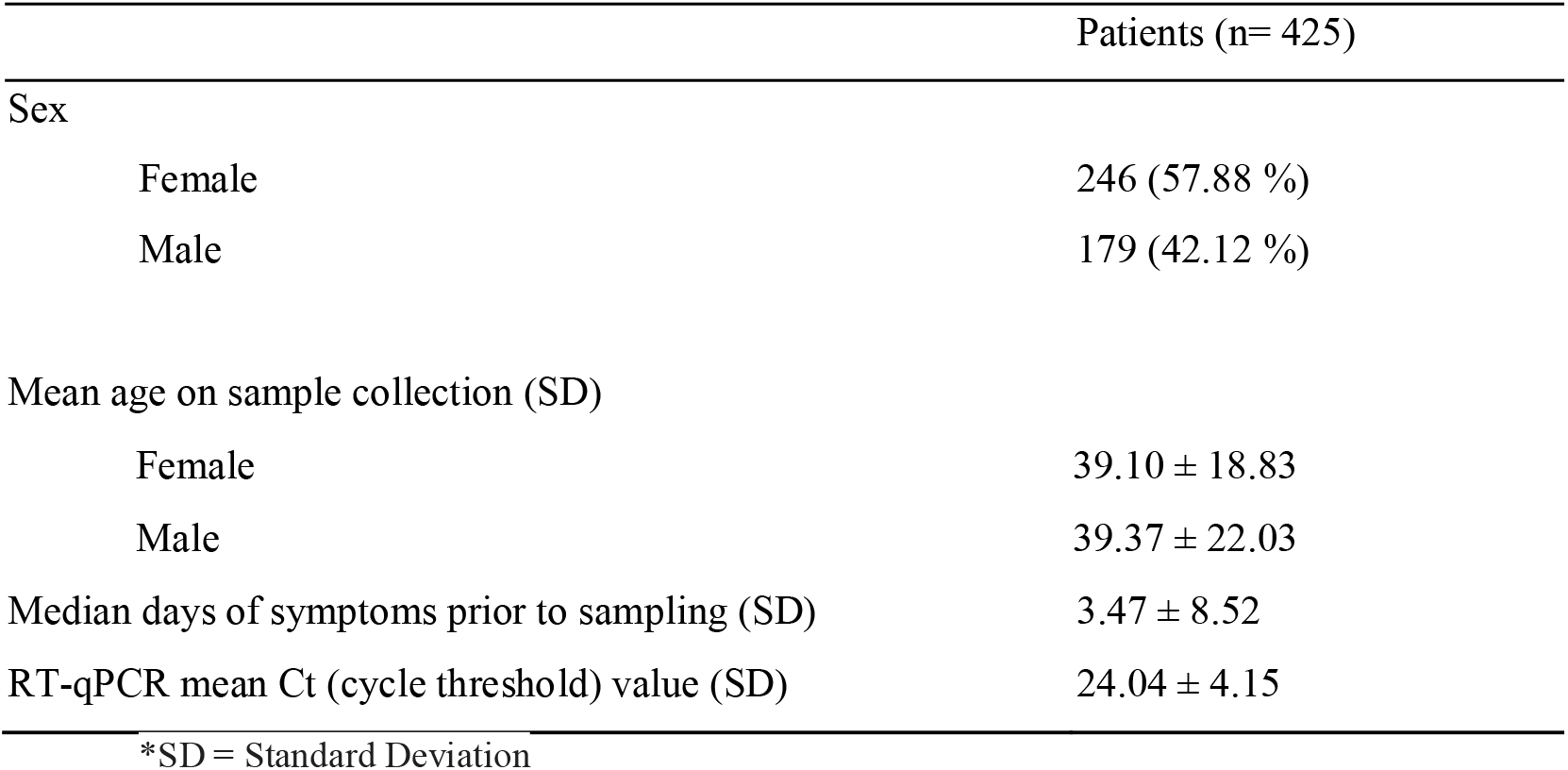
Demographic and laboratory characteristics of Chikungunya virus-infected patients.

Multiplex PCR-tiling amplicon sequencing on MinION allowed the recovery of 425 genomic sequences from CHIKV with a mean genome coverage of 90.98% (range 31.80 to 96.19%) (**Table S1)**. Of the 425 sequences, 14 were recovered from old CHIKV-PCR samples collected in Bahia state during July-August 2015 and stored since. These isolates had a mean genome coverage of 70% (range 31.80 to 92.9 %). The remaining genomes have an associated collection date ranging from April 2019 to June 2022. To better capture the phylogenetic signal, only the sequences with genome coverage over 60% (n=422) were considered for further analysis (discarded sequences are listed in the methods).

All the newly recovered genomes were assembled using Genome Detective software which also classified all of them as belonging to the East, Central, and South African (ECSA) lineage. To investigate the phylogenetic relationship of the new sequences with other Brazilian and non-Brazilian sequences available in public databases, we built a global dataset (n=1,987) composed of 1,565 CHIKV genomes retrieved from GenBank NCBI in addition to 422 sequences from this study. It is noteworthy that the two years of genomic surveillance activities of this study contributed to a more than doubling of the number of CHIKV genomes from Brazil available in the NCBI (by then there were 332 complete sequences) since the virus emerged in the country in 2014 (**Figure 2C**).

**Figure 2.**
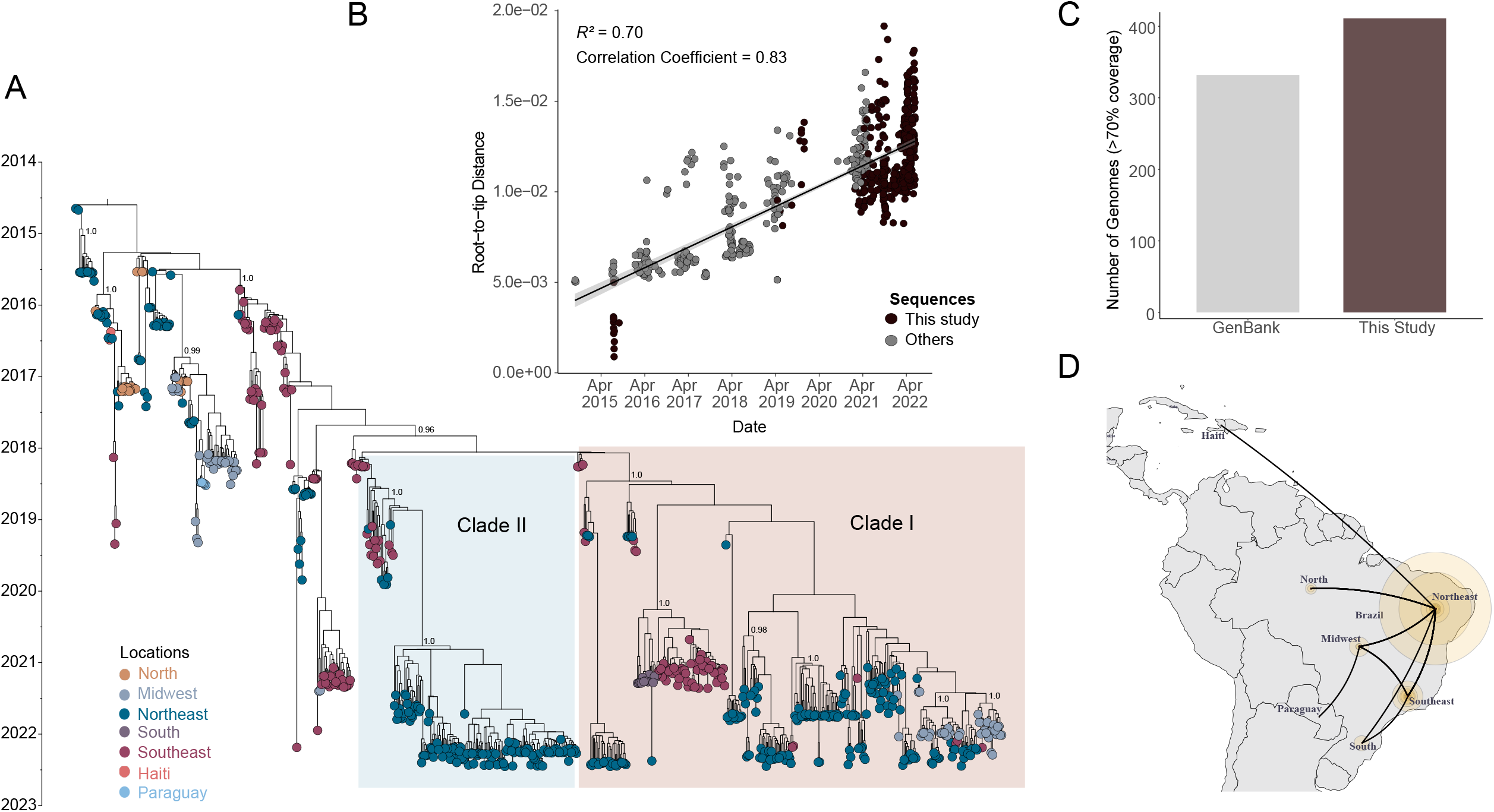
Time-measured phylogeny of CHIKV ECSA lineage in Brazil. A) Maximum Clade Credibility tree reconstructed using 706 sequences from Brazil (in addition to 5 sequences from Paraguay and 2 from Haiti) and a molecular clock approach. Numbers in black show clade posterior probabilities of main nodes. Some posterior probability values were omitted for clarity. Tip colors represent the sampling location. B) Root-to-tip genetic distance regression in a maximum likelihood phylogeny of the CHIKV ECSA lineage (n=713). New sequences are colored in black. C) Number of genomes generated in this study (with >70% genome coverage) compared to the number of Brazilian CHIKV-ESCA sequences available on the GenBank up to 27^th^ Jan. 2023. D). The spatial spread of CHIKV in Brazil estimated under a discrete diffusion model employed in the Bayesian Phylogeographic approach using a dataset with 471 sequences. Size of colored circles was scaled by location posterior support.

### Updated CHIKV phylogeny reveals two distinct emerging subclades

A preliminary Maximum Likelihood (ML) tree was reconstructed using the global dataset that showed all Brazilian sequences grouped in the ECSA clade (**Figure S1**). It can also be observed from the ML tree that most of the new sequences formed two well-distinct derived clades, where it can be noticed that sequences collected in different geographical regions were closely related (see ML tree in Supplemental files). To investigate in more detail the phylogenetic features of these clades within a time-aware evolutionary framework, which can benefit from the increased amount of genomic data obtained in this study, we performed a Bayesian phylogenetic analysis using a down-sampled dataset (n=713) mostly composed of Brazilian sequences.

Root-to-tip genetic distance regression indicated that the down-sampled dataset presented sufficient temporal signal (R^2^=0.70 and correlation coefficient = 0.83) to infer a time-measured phylogeny (**Figure 2B**). Consistent with the ML tree, the inferred Maximum Clade Credibility (MCC) tree also revealed two distinct more derived clades (henceforth clade I and II) formed mainly by 2021-2022 sequences (**Figure 2A**). The Bayesian evolutionary analysis estimated the time of the most recent common ancestor (tMRCA) of clade I to be late January 2018 (95% highest posterior density (HPD): December 2017 and March 2018), while clade II presented a slightly late tMRCA estimated to be early February 2018 (95% HPD: January 2018 and March 2018).

Some composition differences can be noticed between these clades. Clade I comprises sequences from 14 distinct states mostly collected from 2021 to 2022 (n=304) and from northeastern Brazil (62.38%, n=204), with sequences from Sergipe (13.5%, Northeast), Minas Gerais (1.8%), São Paulo (16.2%, Southeast), Goiás (11.6%), Mato Grosso do Sul (0.6%, Midwest), and Paraná (4.3%, South) states uniquely present in this clade (**Figure 2A**). Meanwhile, clade II is mostly composed of sequences collected in 2022 from northeastern states (87.5%, n=147), with a total of 8 states sampled and Piaui state being the most represented (46.42%, n=78) (**Figure 2A**).

A closer look at sequence distribution inside these clades reveals recurrent virus movement between states and regions, with midwestern isolates closely related to isolates from the Northeast and from the state of Minas Gerais (Southeast) in clade I. It can also be noticed in clade I that several distinct CHIKV introductions occurred into the state of Goiás (Midwest) and into northeastern states (Bahia, Rio Grande do Norte, Sergipe, Paraiba, Pernambuco, and Piaui) (**Figure 2A**). Contrarily, the clade I sequence distribution reveals that apparently, only one viral introduction event has happened in the southern state of Paraná, sharing a most recent common ancestor (dated from Jun. 2019 to Jun. 2020, 95% HPD, posterior probability = 1.0) with isolates from the state of São Paulo (Southeast cluster). Similarly, clade II displays viral exchange between states, especially from the Northeast, as indicated by a single well-supported subclade (posterior = 1.0), dated from Jan. 2020 to Dec. 2020 (95% HPD) and dominated by sequences from northeastern states (Piauí, Bahia, Rio Grande do Norte, Paraíba, Pernambuco, Maranhão, and Alagoas) (**Figure 2A)**.

### CHIKV dispersal in Brazil has been mainly seeded by the Northeast

In face of the recurrent virus movement observed across the country, as indicated by our MCC phylogeny, we employed a Bayesian phylogeographic approach to reconstruct the spatial dispersal dynamic of CHIKV in Brazil (closely related sequences from Haiti and Paraguay were included) and to estimate the ancestral locations of clades I and II. The resulting phylogeny kept the topology from the MCC tree shown in figure 2A and revealed the Northeast as a leading source of CHIKV transmission in Brazil, seeding the network of frequent virus exchange among states mainly from the Northeast, Southeast and Midwest, as indicated by the location probability of the 5 early branching events inferred by the discrete phylogeography (**Figure 2D and Figure S2**). Moreover, the ECSA lineage circulating in Brazil extended its transmission network by reaching other countries in the region such as Paraguay and Haiti likely via the Midwest and Northeast of Brazil, respectively. An alternative approach, using a transmission network generated from transition states summarized from the Bayesian phylogeography and centrality metrics, also indicated the Northeast as a source (Source Hub Ratio of 0.66) in the network for the CHIKV spread in the country, where intense interactions are displayed between the Northeast and Southeast (**Figure S3** and **Table S2**). Moreover, the discrete state ancestral reconstruction employed in our Bayesian analysis estimated that both clades I and II might have emerged in the Southeast region with a location probability of 1.0 and with estimated divergence times (clade I: 95% HPD Nov. 2017 – Feb. 2018; clade II: Dec. 2017 – Mar. 2018) comparable to those obtained in the MCC tree inferred using a comprehensive dataset displayed in Figure 2A (**Figure S2**).

These estimates place the divergence time of clades I and II in the period that marks the return of CHIKV-increased transmissions after two main epidemic seasons registered from 2016 to 2017 when a total of more than 565 thousand disease cases were notified in the country (**Figure 1B**). From the time series graph of CHIKF cases, we can see an increase in the incidence rate around early 2018 for the Midwest and North regions. In that same period, clades I and II were estimated to emerge in the Southeast region, which also presented an increased incidence rate. Since then, a seasonal epidemic pattern has been observed in the CHIKV transmission dynamic with incidence peaks being displayed in the first semester of the following years (**Figure 1B**). Consistent with the phylogeography and the transmission network analyses, the CHIKF case time-series plot presents the Northeast region as a major source of virus transmission in the country, as shown by the consecutive incidence peaks registered for that region in the last three years.

### Transition changes outline the evolutionary expansion of the Brazilian ECSA lineage

Despite having close divergence times, clades I and II have different sequence compositions that likely explain their apparent branching distance. While sequences from clades I and II were obtained from patients with similar median age, a significant difference (p<0.05) was observed between the median RT-qPCR cycle threshold value of the samples of each clade (**Figure 3A**). Such a difference, however, might have been affected by an imbalance present in the dataset used for comparison since clade I has almost twice as many sequences compared to clade II.

**Figure 3.**
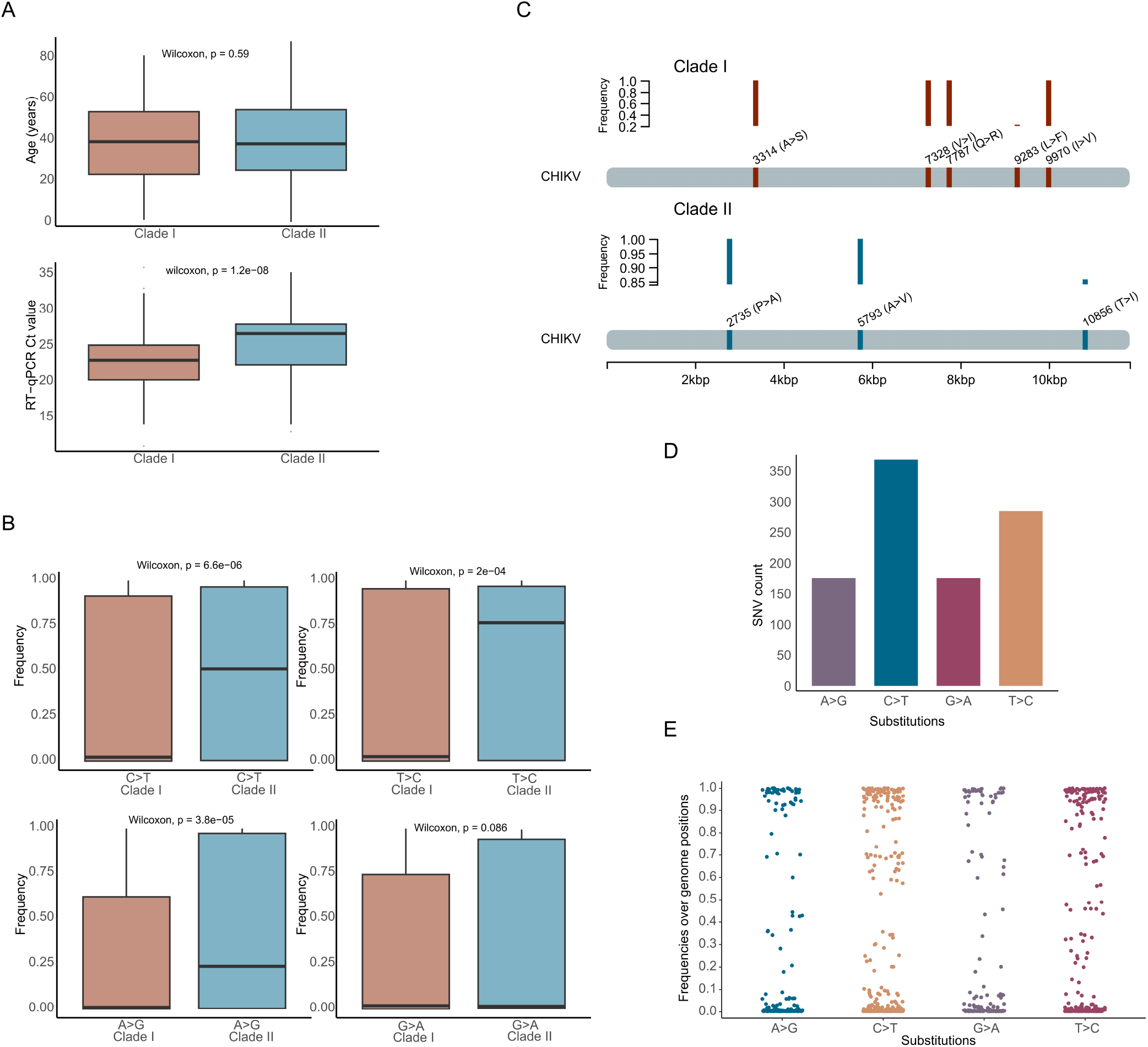
Genetic and demographic aspects of the newly generated CHIKV sequences. A) Boxplot of the patient’s age and samples’ cycle threshold value distributions by clades I and II. Mann-Whitney U test with a significance level alpha = 0.05. B) Boxplot of the frequency distributions for each class of single nucleotide variation (SNV) identified in the genomes from clades I and II. Mann-Whitney U test with a significance level alpha = 0.05. C) Genome positions and frequency of the non-synonymous substitutions uniquely identified in sequences from clades I and II. D) Bar plot of the absolute number of CHIKV genome mutated positions for each class of single nucleotide variation (SNV) identified in the alignment of the new CHIKV sequences (n=422). E) Scatterplot of the frequency distributions for each class of single nucleotide variation (SNV) identified in the alignment of the new CHIKV sequences (n=422). Each dot represents a distinct mutated genome position.

We also assessed these clades as to which selective regime they are likely to be subject to. The results from the BUSTED analysis provided evidence that the envelope gene experienced positive diversifying selection in both clades I (p=1.774e-8) and II (p=1.840e-11) (**Table S3**). For comparison, we employed a second method, MEME, that identified 12 sites under positive selection for clade I, whereas 13 sites were identified in clade II. Of these sites, 7 are shared by both clades, while exclusive positive selected sites were reported for each clade (**Table S4)**. These results suggest the active status of these clades and consequently provide evidence of the continuous evolutionary expansion of the Brazilian ECSA lineage.

Separate sequence alignments representative of each clade revealed a significant difference between the median frequency of 3 classes of single nucleotide variation (SNV) identified in the clades. Clade II presented a higher median frequency of SNV of type C>T, A>G, and T>C transitions (**Figure 3B**). By comparing the mutational profile of the two clades, we identified 27 SNVs exclusively present and shared by sequences from clade II, with three of them being non-synonymous substitutions leading putatively to amino acid change such as a T288I substitution in the E1 protein gene (**Table 3 and Figure 3C**). The other two non-synonymous substitutions were identified in two nonstructural proteins (nsP2-P352A and nsP4-A43V). Sequences from Clade I, on the other hand, presented 13 exclusive SNVs, of which six are non-synonymous mutations present in two nonstructural proteins (nsP2 and nsP4) and three structural proteins genes (capsid, 6k and E2) (**Table 3 and Figure 3C**). The capsid gene sequence from this clade presented two contiguous transition substitutions in the same codon (7787-7788) resulting in a Q74R change. In an alignment analysis of all 422 new sequences, we found that among the total of CHIKV genome mutated positions the majority consisted of type C>T transitions (29.5%, n= 368) followed by T>C substitutions (22.9%, n= 285) (**Figure 3D**). These transitions along with A>G and G>A transitions displayed a comparable frequency distribution across the sequence dataset, where mutations at several positions were identified in all new sequences (**Figure 3E**). Due to the possibility of these substitutions resulting from sequencing errors, we performed the same comparative analysis on a different dataset containing sequences that form a subclade (from the South region) in clade I and that were generated by a different sequencing technology (Illumina) and a different research group (data in Supplemental files). We observed the same pattern of increased frequency (26.8%) of C>T transitions among those southern sequences, which suggested the observed increased transition changes in clades I and II were likely derived from mutation rather than sequencing artifacts.

**Table 3.**
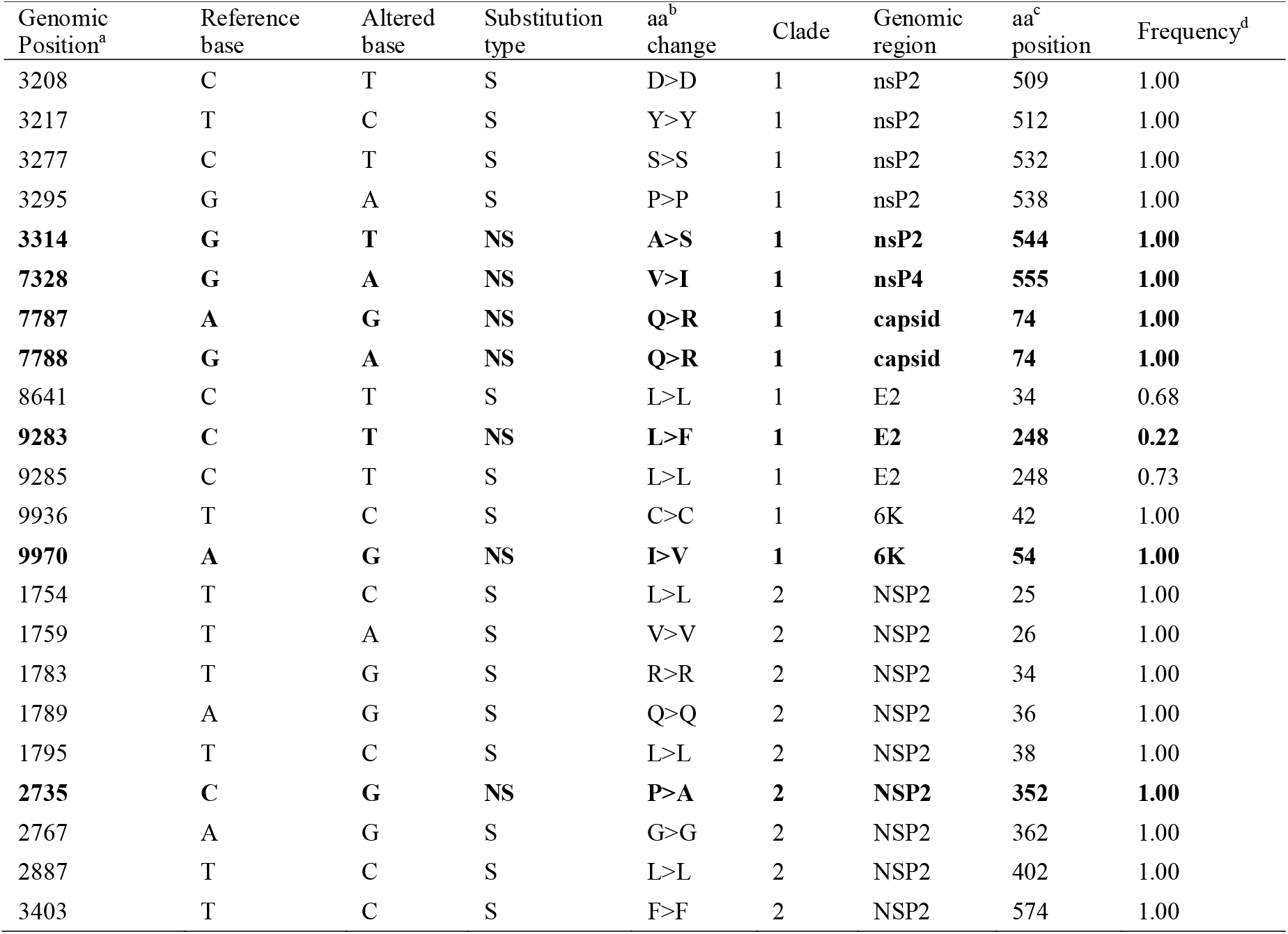
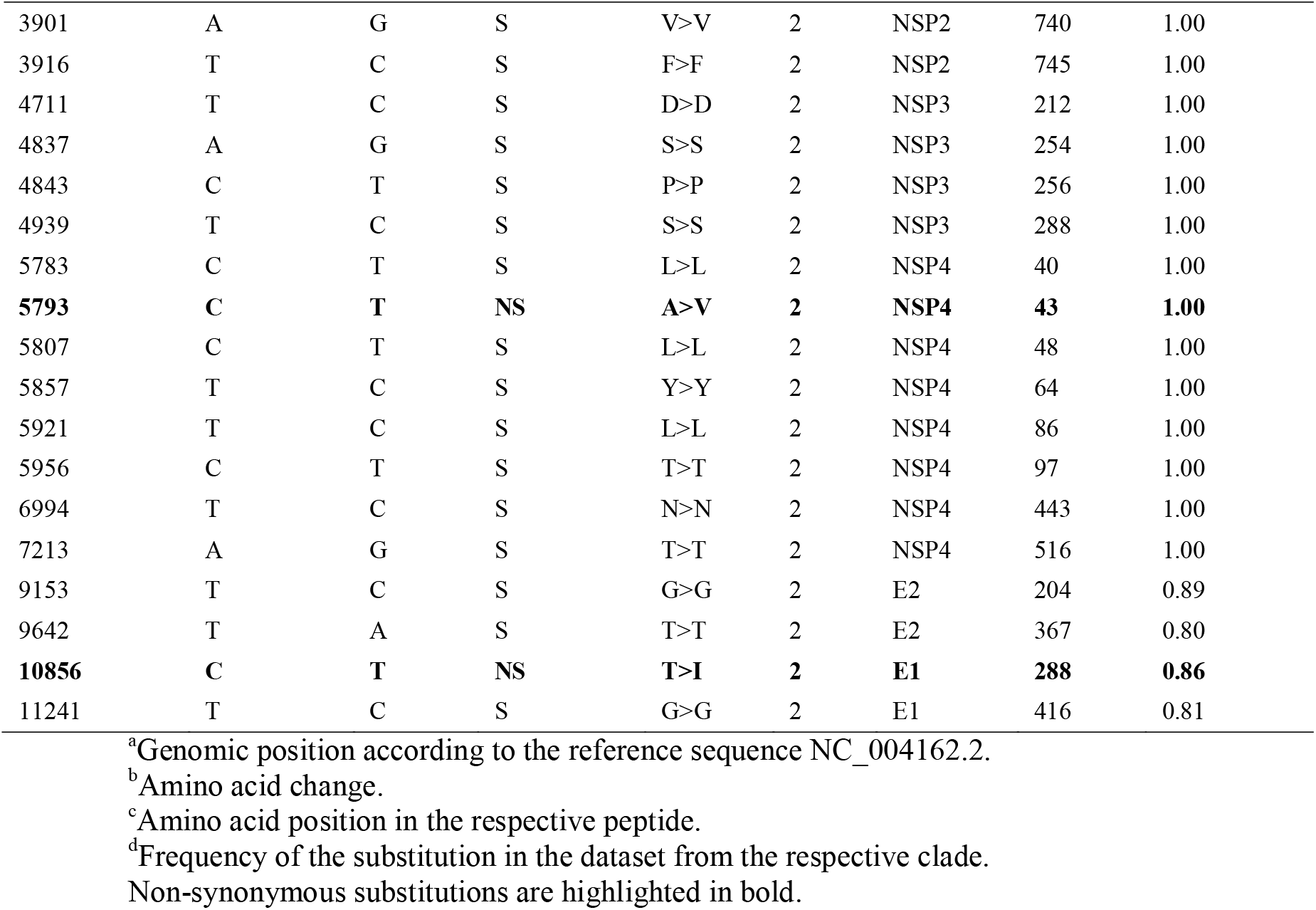
List of single nucleotide variations (SNV) uniquely identified for clades I and II.

## Discussion

Advances in sequencing technology and bioinformatics tools have contributed to an increased understanding of the genetic diversity and transmission dynamics of emerging viruses causing epidemics ^25^. In this study, we increased the number of CHIKV genomes deposited in the NCBI by generating 422 genomes that cover 12 Brazilian states over the years 2021-2022. By combining genomic and demographic data this effort has provided a comprehensive overview of CHIKV ECSA phylogeny in Brazil which summarizes and updates previous phylogenies from other studies of localized outbreaks ^10,12,14,26,27^. The updated and metadata-enriched dataset allowed us not only to describe the genetic diversity of circulating CHIKV variants but also to describe branching patterns observed in the updated phylogeny and infer the divergence time and likely location of emerging distinct subclades from the Brazilian lineage.

Following the arrival of ECSA lineage in the Northeast region, a number of other studies have predicted and reported the establishment and spread of this lineage to the rest of the country ^10–12,14,17,26–29^. The Bayesian phylogeographic approach employed in this study has revealed the main routes of dispersion of CHIKV in the country. After being introduced in the Northeast region, the ECSA lineage dispersed towards several states from all five regions in Brazil. Both the Bayesian phylogeographic and transmission network approaches indicated the Northeast region has been acting as the leading hub of virus spread towards other regions in the country, including by forming an intense virus exchange network with the Southeast region. From the Southeast, the virus dispersed to the southern state of Paraná (posterior probability of 0.96) with the time of the most recent common ancestor shared with samples from São Paulo estimated to be January 2020. In addition to spread in Brazil, CHIKV has also extended its circulation to other countries such as Paraguay and Haiti. CHIKV circulation in Paraguay and Haiti has been previously associated with transmission events originating from Brazil through international viral exchange likely mediated by human movement ^30–32^. These results evidence the lineage’s potential expansion across Latin America, despite limited information available about this lineage in the region as evidenced by the lack of CHIKV ECSA sequences from other Latin American countries.

Our time-measured phylogeny corroborated previous studies and revealed emerging branching patterns, mainly represented by two well-supported distinct subclades named clade I and II. Both these clades were estimated to emerge in the Southeast region around the first months of 2018. At the end of that year, the Southeast region accounted for more than 65 thousand reported cases. Moreover, differences in sequence composition were observed between these clades, with clade I being more diverse as it comprises sequences from four different regions (Northeast, Midwest, Southeast and South) collected in the years 2021— 2022, while clade II contains mostly sequences from northeastern states collected in 2022. This difference in the clades sequence composition profile might change as the lineage continue to expand into the country. The observed differences in geographic diversity between clades might reflect distinct transmission networks underlying the divergence and expansion of these subclades.

Different viral lineages might be under distinct evolutionary pressure that together with the emergence and selection of mutations can drive viral adaption to a particular environment ^33,34^. It has been argued that two different CHIKV lineages, IOL and Asian, have undergone different evolutionary trajectories leading to different vector adaptative potentials ^35^. Here, we used the ratio of non-synonymous (*dN*) to synonymous (*dS*) nucleotide substitutions in the CHIKV envelope gene to assess the selective regime to which clades I and II might be subjected. Our analysis revealed that both clades have experienced positive diversifying selection. Such evolutionary pressure might be driven by the host antiviral immune response, as the viral envelope protein is a target for neutralizing antibodies ^36,37^. Mutations in the envelope proteins have been implicated in the increased adaptation and transmission of the CHIKV IOL lineage in *Aedes albopictus* mosquitoes ^22,38^.

A higher ratio of nonsynonymous substitutions is observed under a positive selection regime promoted by virus-host interactions ^39^. Since SNVs continue to arise in RNA virus populations mainly driven by errors made by the virus replication complex that lead to genetic diversity, we compared the mutational profiles of clades I and II ^40^. We identified several SNVs across nonstructural and structural protein genes that were exclusive to each clade. Clade II presented more SNVs (n=27) than clade I (n=13), of which three are non-synonymous substitutions (E1-T288I; nsP2-P352A, and nsP4-A43V). Literature research revealed that E1-T288I change was previously identified in a 2017 sequence from Iran and also in CHIKV sequences collected in 2016 from infected cancer patients in Rio de Janeiro, Brazil ^41,42^. The nsP2-P352A substitution was also present in sequences collected between 2016 and 2017 in Rio de Janeiro ^14,43^. In turn, clade I sequences contain six non-synonymous mutations across nonstructural (nsP2 and nsP4) and structural protein genes (capsid, 6k and E2). The nsP4-V555I change was previously detected in sequences from Thailand in 2008-2009 ^44^. The E2-L248F from clade I has been reported in Asian lineage sequences from Colombia (2014-2015) and Philippines (2012)^45,46^. Isolates from Thailand, Indonesia, Lao PDR, Cameroon and India also presented the 6K-I54V mutation observed in clade I^44,47–50^. Despite the detection of these mutations in different countries (indicating homoplasy) by other previous studies, there is no information about the functional impact of such substitutions on CHIKV fitness, thus warranting further experimental studies to elucidate the potential effects of SNVs on lineage-specific evolutionary adaptation. It has been argued that not only non-synonymous mutations have the potential to promote adaptive changes but also synonymous mutations can lead to changes in the viral RNA that can drive differential viral gene expression^51^.

Mutational analysis of the 422 sequences generated in this study revealed a higher amount of C>T and T>C transitions followed by A>G and G>A substitutions in the CHIKV genome and several genome positions presenting these transitions with higher frequency across all new sequences. Moreover, Clade II has a significantly higher median frequency of C>T, A>G, and T>C transitions compared to clade I. Although this study cannot experimentally establish the significance of these transitions for CHIKV evolutionary adaptation, other studies have associated this mutational pattern with the action of host antiviral immune repose mediated by AID/APOBEC and ADAR families of deaminases^52,53^. These enzymes are part of the interferon-stimulated innate immune response and promote viral genome transitions mutations by catalyzing the deamination of adenosine to inosine to cause A>G/T>C (by ADAR) substitutions or deamination of cytosine to uracil that leads to C>T/G>A (by AID/APOBEC) mutations^54^. This RNA editing process has been experimentally observed targeting specific viral sequences of SARS-CoV-2 to produce C>T transitions and increasing viral replication in Caco-2 cells, thus promoting viral increased fitness and adaptative evolution^55^. However, specific information about the effect of these RNA editing processes on the CHIKV genome remains elusive, although the APOBEC3A gene has been observed up-regulated in the expression profile of CHIKV-infected patients^56^. Despite arguments that these transitions happen mainly in phylogenetically uninformative sites, the available evidence indicates that RNA editing processes might act as a significant driver of viral sequence diversity and evolutionary adaptation through the introduction of nucleotide changes^53,57^.

Recurring CHIKF epidemics, as indicated by the seasonal peak patterns displayed in the case time-series plot, are evidence that the virus is endemic in Brazil. Human mobility, population immunity, vector suitability, vegetation coverage, site socioeconomic status, and viral sequence variation are factors considered to mediate the dispersal of CHIKV in Brazil^26,58,59^. Although we did not find the E1:226V *Aedes Albopictus-*adaptive mutations in the Brazilian sequences, the high abundance in the region of widely spread competent vectors, such as *A. aegypti* and *Ae. Albopictus*, together with favorable climatic and social conditions in large urban centers create conditions that modify the adaptive landscape of CHIKV, which in turn can allow the continued expansion of the ECSA lineage in the country with a resultant increased impact on public health^23,60,61^. Therefore, public health measures should be undertaken to ensure continuous genomic surveillance of circulating CHIKV variants which can help to identify viral transmission routes where focused vector control strategies could be employed to reduce the risk of recurring CHIKF epidemics.

### Limitations

Although our study presented the results of a Bayesian phylogeographic and mutational profile analysis performed on 422 new sequences collected from 12 Brazilian states, not all states were evenly represented in our dataset, which might limit our estimates relative to divergence time and ancestral location reconstructions, prompting careful interpretation of the results presented here. Ongoing sequencing efforts across the country could reduce this disparity in the future. Moreover, although single nucleotide substitutions identified among the new sequences offer insights into the evolutionary dynamics of CHIKV in Brazil, further functional studies need to be undertaken to elucidate the actual adaptive effect of these mutations.

## Data Availability

New sequences have been deposited in GenBank. Input files used for the phylogenetic and mutational profile analyses will be available at the GitHub repository https://github.com/genomicsurveillance. All data produced in the present study are available upon reasonable request to the authors.

## Acknowledgements

This work was supported by The Pan American Health Organization (PAHO/WHO), the Brazilian Ministry of Health grant SCON2021-00180 (Coordenação Geral de Laboratório de Saúde Pública-CGLAB and Coordenação Geral de Vigilância de Arboviroses-CGARB), the National Institutes of Health USA grant U01 AI151698 for the United World Arbovirus Research Network (UWARN). We thank the State Health Secretariats and the epidemiological surveillance services of Brazilian states and municipalities for all the support provided. JX is supported by Coordenação de Aperfeiçoamento de Pessoal de Nível Superior-Brasil (CAPES)-Finance Code 001. MG is funded by PON “Ricerca e Innovazione” 2014-2020.

## Author Contributions

**Conception and Design:** L.C.J.A., M.G., V.F., J.X.; **Investigations**: J.X., M.G., V.F., L.C.J.A., C.O., H.F., T.A., N.G., S.T., M.E., E.S., E.V.S., L.M., E.C., M.L., D.L.R., A.F.M., A.V., C.N.D.S., C.Z., C.S., D.C., F.C., G.B., H.B., I.G., I.R., I.C.S., J.B.S., G.A., J.L., J.A., J.P., K.G.L., L.B.S., L.A.F.S., M.G.A., M.L., M.B.F, M.C.S.U.Z., M.P., N.F., N.M., R.V.C., K.S.T., S.K.H., S.N.S., T.A., T.C., V.N., G.C.P., F.I., A.L.S.M., F.P., W.G.C., N.A., M.C.B.C., T.R., C.A.S., L.S., V.S., L.A.P., L.G.L.N., H.F., B.V., D.T., D.F.L.N, R.H.S., J.M., I.N.R., L.D., G.G.C.L., A.L., J.P.M.N., C.C.M.G., E.C.O., R.J., R.K., A.M.B.F.; **Data Curation**: J.X., M.G., V.F., L.C.J.A; **Formal Analysis**: J.X., M.G., V.F., L.C.J.A.; **Writing–Original Draft Preparation**: J.X., M.G., L.C.J.A.; **Revision:** J.X., M.G., V.F., L.C.J.A.; **Resources**: J.M.R., M.A., W.N.A., A.R., R.F.C.S., C.F.C.A., W.C.V.V., P.R., M.G.J., J.N.W., R.S., E.L.N.M., P.EA.S., H.C.F.F., C.F., N.F.O.M., L.C.V.F., A.M.B.F., L.C.J.A.

## Data and code availability

New sequences have been deposited in GenBank under accession numbers [available soon]. Input files used for the phylogenetic and mutational profile analyses will be available at the GitHub repository https://github.com/genomicsurveillance.

## Methods

### Sample collection

Residual samples (serum or plasma) were obtained from the epidemiological surveillance routine of the Brazilian Central Public Health Laboratories (LACEN) from different states (Alagoas, Bahia, Goiás, Paraíba, Paraná, Pernambuco, Piauí, Maranhão, Minas Gerais, Mato Grosso do Sul, Rio Grande do Norte, and Sergipe). These samples were submitted to nucleic acid purification using the MagMax Viral RNA Mini kit (Thermo Fischer Scientific), following the manufacturer’s recommendations, and were previously screened by each LACEN. CHIKV RT-qPCR positive samples were selected for sequencing based on the cycle threshold value ≤ 30 and the availability of demographic metadata such as sex, age, and municipality of residency. These demographic patient data were provided by LACENs and were collected through a questionnaire filled out at local health care services.

### Ethical statement

The project was approved by the Pan American World Health Organization (PAHO) and the Brazilian Ministry of Health (MoH) as part of the arboviral genomic surveillance efforts within the terms of Resolution 510/2016 of CONEP (Comissão Nacional de Ética em Pesquisa, Ministério da Saúde; National Ethical Committee for Research, Ministry of Health). This authorizes the use of clinical samples collected in the Brazilian Central Public Health Laboratories to accelerate knowledge building and contribute to surveillance and outbreak response. The study protocol was reviewed and approved by Research Ethics Committee of the Universidade Federal de Minas Gerais with approval No. 32912820.6.1001.5149. Personally identifying information were de-identified in the datasets and tables in a way that minimizes the risk of unintended disclosure of identity of individuals and information about them.

### cDNA synthesis and whole genome sequencing

Extracted RNA from positive CHIKV samples were provided by collaborating LACENs and submitted to cDNA synthesis and PCR, using a sequencing protocol based on multiplex PCR tiling amplicon approach design for MinION nanopore sequencing^62^. All reactions were performed at biosafety level 2 facilities and using no template controls. PCR products were purified using 1x AMpure beads Beckman Coulter, UK) and quantified using Qubit 3.0 instrument (Life Technologies) and the Qubit dsDNA High Sensitivity assay. DNA library preparation was performed on all amplified samples using the Ligation Sequencing Kit (Oxford Nanopore Technologies). Individual samples were barcoded using the Native Barcoding Kit (NBD104, Oxford Nanopore Technologies, Oxford, UK). Sequencing library was loaded onto a R9.4 flow cell and data were collected for up to 48 sequencing hours.

### Generation of consensus sequences

Basecalling of raw FAST5 files and demultiplex of barcodes were performed using the software Guppy (https://github.com/nanoporetech). Consensus sequences were generated by a hybrid assembling approach implemented on Genome Detective (https://www.genomedetective.com/)^63^.

### Phylogenetic reconstruction

We used MAFFT to align 422 new sequences (with coverage over >60% according to Thézé et al. ^66^, samples 736.22_RED, FS0116, and FS0132 were discarded) in addition to 1,565 CHIKV whole genome sequences publicly available in NCBI up to August 2022, forming a global dataset (n=1,987) that includes all lineages. This global dataset was used to infer a Maximum Likelihood (ML) phylogeny using the IQ-TREE 2.1.1 software^64,65^. Statistical support for tree nodes was estimated using the ultrafast bootstrap (UFBoot) feature implemented in IQ-TREE with 1,000 replicates. We then used the ML tree from the global dataset to extract the Brazilian ECSA clade and use it to form a second dataset (total n=713, 706 Brazilian sequences, 2 from Haiti, and 5 from Paraguay; sequences with genome coverage >60% according to Thézé et al. ^66^) which was used to infer a time-scaled phylogeny using BEAST v1.10.4. First, we investigated the temporal signal regressing root-to-tip genetic distances from this ML tree against sample collection dates using TempEst v.1.5.1^67^. Secondly, we employed a stringent model selection analysis using both path-sampling (PS) and stepping-stone (SS) procedures to estimate the most appropriate molecular clock model for the Bayesian phylogenetic analysis^68^. For the Bayesian analysis, the uncorrelated relaxed molecular clock was chosen as indicated by estimating marginal likelihoods, also employing the HKY+G4 nucleotide substitution model, and the nonparametric Bayesian Skyline coalescent model. We combined two independent runs of 200 million states each^69^. The convergence of MCMC chains was checked using Tracer^70^. Maximum clade credibility (MCC) trees were summarized using TreeAnnotator after discarding 10% as burn-in.

CHIKV ECSA lineage movements across Brazil were investigated using the Bayesian phylogeographic approach with a discrete trait phylogenetic model. A trait file was used to discretize sequences sapling location by five Brazilian regions (North, Northeast, Southeast, South and Midwest). For this analysis, we downsampled our Brazilian ECSA clade to a dataset containing 471 sequences to maximize the temporal signal in the dataset. MCMC analyses were performed in BEAST v1.10.4, running in duplicate for 200 million interactions and sampling every 20,000 steps in the chain. Convergence for each run was assessed in Tracer. MCC trees for each run were summarized using TreeAnnotator after discarding the initial 10% as burn-in. Finally, we used SPREAD 4 tool to map spatiotemporal information embedded in the MCC trees^71^.

### Transmission network analysis

A transmission network was reconstructed, using the StrainHub tool, from transition states summarized from the Bayesian phylogeography^72^. Centrality metrics on the tree nodes were also estimated for the network.

### Comparative mutational analysis

For comparative mutational analysis, we assembled separate alignment datasets for each subclade (clade I = 327; clade II = 168) and for all new sequences (n=422). The sequence datasets were compared against the NCBI reference strain NC_004162.2 using MALVIRUS^73^. We filtered and selected substitutions with an occurrence frequency of 100% across the whole dataset and substitutions with a frequency above 60% across the envelope genes E1 and E2.

### Selection pressure analysis

Since the ratio of non-synonymous (*dN*) to synonymous (*dS*) nucleotide substitutions can be used to study selection pressure on genomic sequences, we used the HYPHY software package that employs statistical methods that estimate the *dN/dS* to detect diversifying selection^74^. For that we performed an alignment of the envelope gene sequences of 720 Brazilian isolates and used BUSTED (restricting the analysis to each subclade I or II), an alignment-wide method implemented in HYPHY, that aims to detect evidence of episodic diversifying selection^75^. For comparison, we also used a different method, called MEME, a site-level approach that aims to detect evidence of both pervasive and episodic diversifying selection at individual sites (also restricting the analysis to each subclade I or II) ^76^.

### Epidemic curves from Chikungunya cases reported in Brazil

Data of weekly notified and laboratory-confirmed cases of infection by CHIKV in Brazil from 2014 to 2022 were supplied by the Brazilian Ministry of Health (BrMoH)(Ministério da Saúde, 2022). These data were used to calculate incidence and to plot time series charts.

