## Supplemental files for "Increased interregional virus exchange and nucleotide diversity outline the expansion of the chikungunya virus ECSA lineage in Brazil"

### Supplemental figures legends

**Figure S1. Global phylogeny of CHIKV lineages.** Maximum Likelihood (ML) phylogeny reconstructed using 1,987 CHIVK sequences, including the new genomes and others available on GenBank. New Brazilian genomes are indicated by blue branches in the Brazilian ECSA clade. Numbers are clade ultrafast bootstrap support values.

**Figure S2**. **Ancestral locations of viruses from the CHIKV ECSA lineage circulating in Brazil**. Maximum Clade Credibility tree reconstructed under a Bayesian phylogeography in discrete space using sequences (total n=471) from Brazil, Paraguay and Haiti**.** Location probabilities of clades I and II as well as of main early branches estimated to be originated in the Northeast of Brazil are represented by numbers placed near nodes. Location probabilities of other clades were omitted for clarity.

**Figure S3**. **Transmission network of CHIKV in Brazil**. The network was inferred from the transitions between geographic states from the Bayesian phylogeography under a discrete space model. Arrows indicate directionality and their thickness indicates transition frequencies. Node size was scaled by source hub ratio values estimated for each location. Colors are arbitrary.

**Figure S1. Global phylogeny of CHIKV lineages**

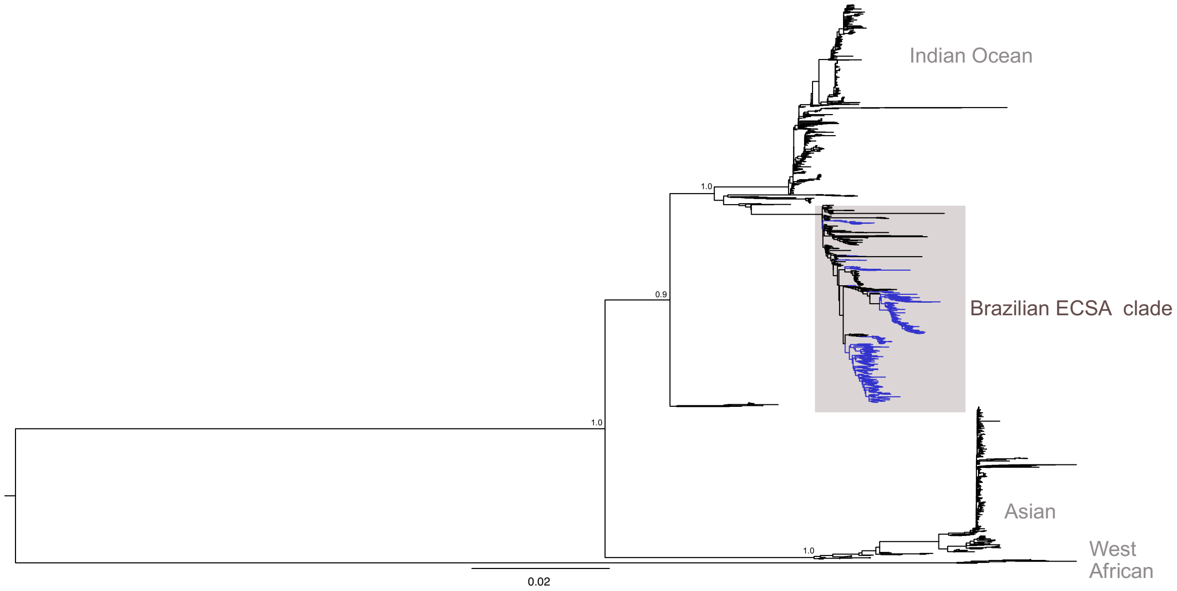

**Figure S2**. **Ancestral locations of viruses from the CHIKV ECSA lineage circulating in Brazil**.

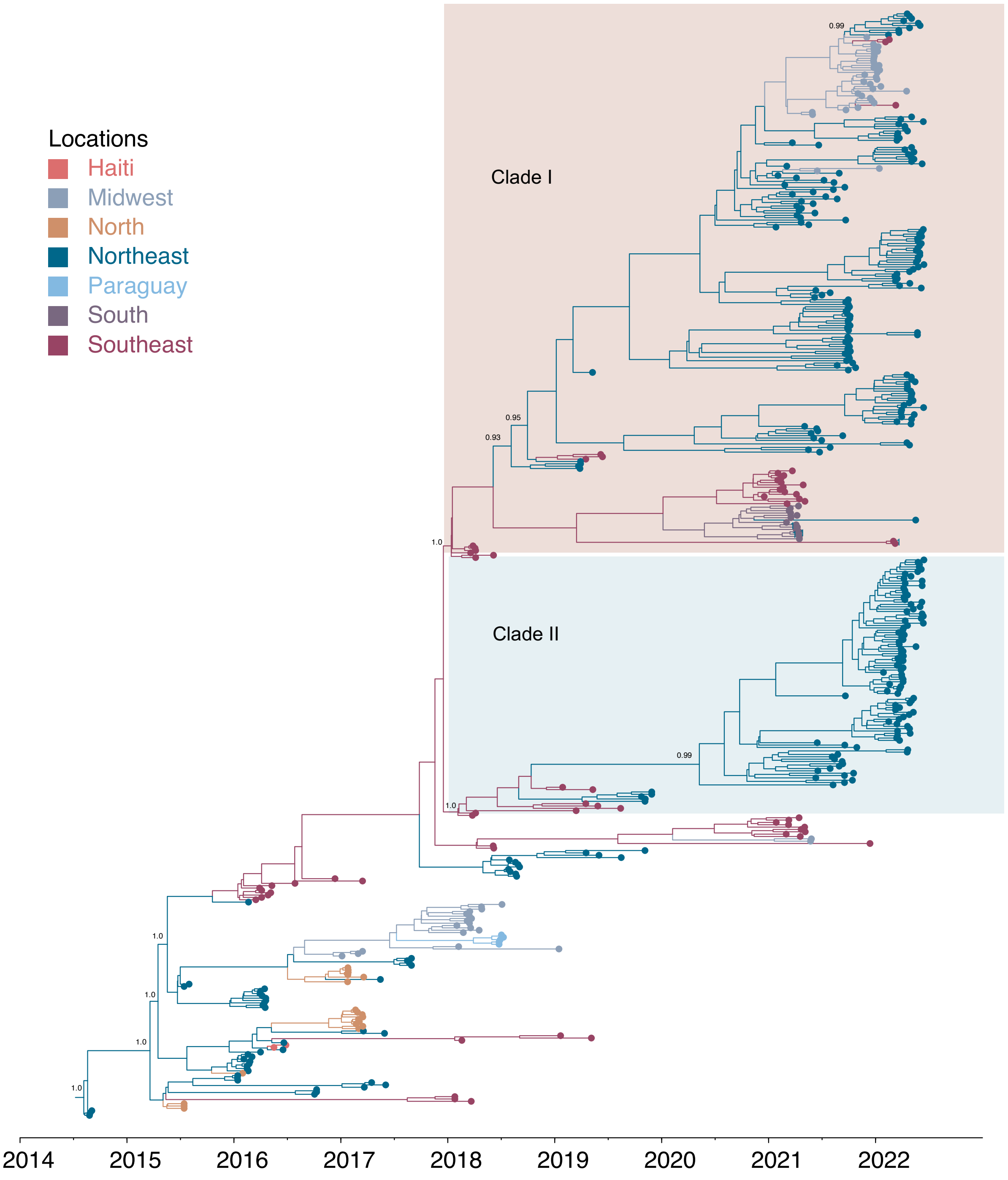

**Figure S3. Transmission network of CHIKV in Brazil.**

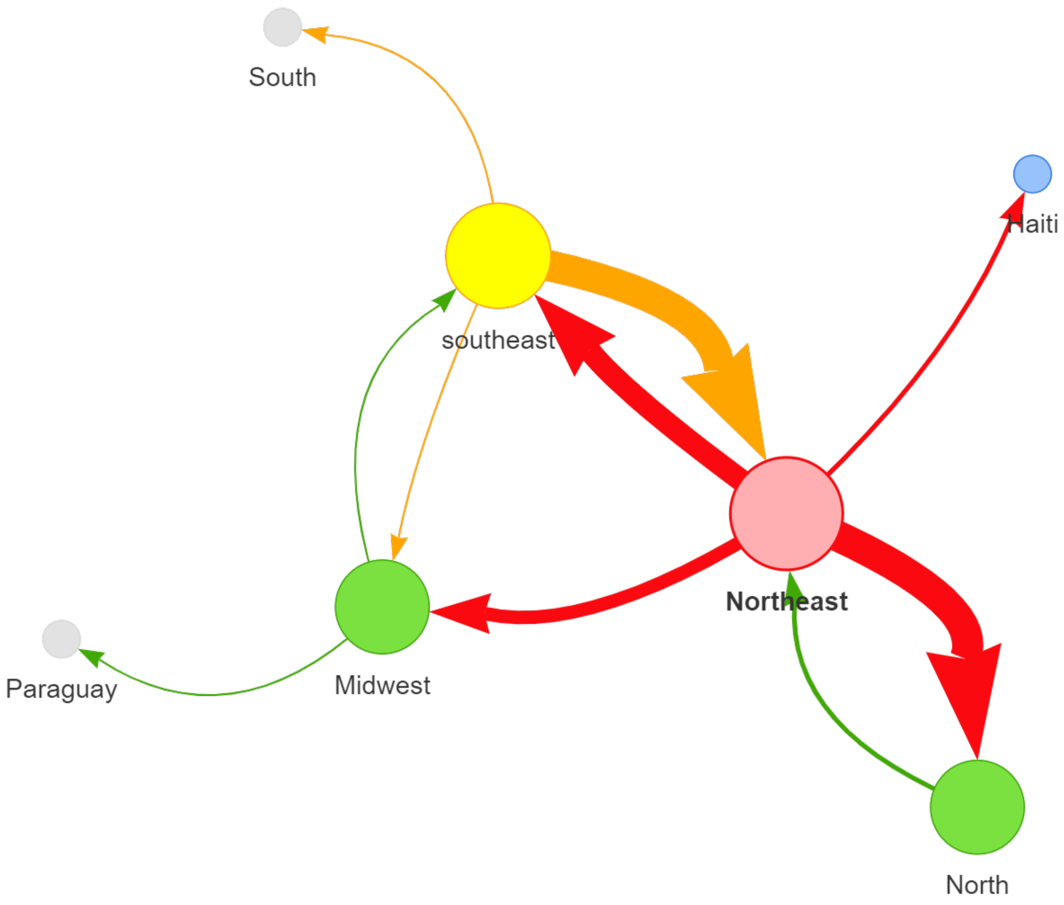

**Table S1. Sample information and sequencing statistics.**

| ID-sequence | Barcode | Lab ID | Ct | State | Municipality | Collection date | Sex | Reads | Coverage | Depth of Coverage | Identity NT | Identity AA |
|---|---|---|---|---|---|---|---|---|---|---|---|---|
| CHIKV\|NB06\|ARB02\|AL\|2021-08 | NB06 | ARB 2 | 21.92 | AL | BARRA DE SAO MIGUEL | 8-2021 | M | 8179.0 | 94.3 | 816.7 | 96.8 | 98.3 |
| CHIKV\|NB07\|ARB08\|AL\|2021-09 | NB07 | ARB 8 | 15.71 | AL | DELMIRO GOUVEIA | 9-2021 | F | 56805.0 | 94.1 | 3498.3 | 96.6 | 98.0 |
| CHIKV\|NB08\|ARB09\|AL\|2021-10 | NB08 | ARB 9 | 21.92 | AL | UNIAO DOS PALMARES | 10-2021 | F | 3038.0 | 92.9 | 226.8 | 96.8 | 98.3 |
| CHIKV\|NB02\|NIH-01\|BA\|2019-04 | NB02 | NIH-01/966 | 27.14 | BA | FEIRA DE SANTANA | 4-2019 | M | 132760.0 | 94.5 | 7528.6 | 96.6 | 98.1 |
| CHIKV\|NB03\|NIH-02\|BA\|2019-06 | NB03 | NIH-02/1435 | 26.52 | BA | FEIRA DE SANTANA | 6-2019 | M | 76168.0 | 94.6 | 5412.9 | 96.7 | 98.3 |
| CHIKV\|NB04\|NIH-03\|BA\|2019-05 | NB04 | NIH-03/1524 | 18.10 | BA | FEIRA DE SANTANA | 5-2019 | F | 2382.0 | 92.7 | 156.6 | 96.9 | 98.3 |
| CHIKV\|NB05\|NIH-04\|BA\|2019-08 | NB05 | NIH-04/1801 | 21.32 | BA | FEIRA DE SANTANA | 8-2019 | M | 88238.0 | 94.6 | 6633.4 | 96.6 | 98.3 |
| CHIKV\|NB09\|BA-01\|2021-02 | NB09 | BA-01 | 14.80 | BA | BARREIRAS | 2-2021 | M | 28095.0 | 94.3 | 3298.8 | 96.8 | 98.3 |
| CHIKV\|NB10\|BA-02\|2021-04 | NB10 | BA-02 | 15.10 | BA | ITAGUACU DA BAHIA | 4-2021 | M | 22200.0 | 94.8 | 2405.0 | 96.8 | 98.3 |
| CHIKV\|NB11\|BA-03\|2021-06 | NB11 | BA-03 | 15.90 | BA | FEIRA DE SANTANA | 6-2021 | M | 48701.0 | 94.2 | 8877.8 | 96.8 | 98.3 |
| CHIKV\|NB12\|BA-04\|2021-03 | NB12 | BA-04 | 18.30 | BA | BARREIRAS | 3-2021 | F | 72292.0 | 94.1 | 5770.4 | 96.8 | 98.2 |
| CHIKV\|NB13\|BA-05\|2021-02 | NB13 | BA-05 | 18.80 | BA | BARREIRAS | 2-2021 | F | 46042.0 | 94.5 | 3880.8 | 96.7 | 98.3 |
| CHIKV\|NB14\|BA-06\|2021-09 | NB14 | BA-06 | 19.10 | BA | SAPEACU | 9-2021 | F | 78195.0 | 94.5 | 6739.6 | 96.7 | 98.3 |
| CHIKV\|NB15\|BA-08\|2021-03 | NB15 | BA-08 | 20.40 | BA | BARREIRAS | 3-2021 | M | 70997.0 | 94.5 | 6115.9 | 96.8 | 98.3 |
| CHIKV\|NB16\|BA-09\|2021-04 | NB16 | BA-09 | 21.00 | BA | BARREIRAS | 4-2021 | F | 97995.0 | 94.4 | 6624.4 | 96.5 | 98.2 |
| CHIKV\|NB17\|BA-10\|2021-04 | NB17 | BA-10 | 21.20 | BA | BARREIRAS | 4-2021 | F | 63373.0 | 94.2 | 4977.7 | 96.7 | 98.3 |
| CHIKV\|NB18\|BA-12\|2021-04 | NB18 | BA-12 | 22.10 | BA | IRECE | 4-2021 | M | 38035.0 | 94.5 | 3366.7 | 96.6 | 98.2 |
| CHIKV\|NB19\|BA-15\|2021-03 | NB19 | BA-15 | 22.80 | BA | BARREIRAS | 3-2021 | M | 68865.0 | 94.2 | 5050.5 | 96.8 | 98.3 |
| CHIKV\|NB20\|BA-18\|2021-07 | NB20 | BA-18 | 23.50 | BA | NOVO TRIUNFO | 7-2021 | F | 26719.0 | 94.1 | 2002.1 | 96.7 | 98.3 |
| CHIKV\|NB21\|BA-19\|2021-08 | NB21 | BA-19 | 23.50 | BA | POJUCA | 8-2021 | F | 59248.0 | 94.8 | 5119.6 | 96.7 | 98.2 |
| CHIKV\|NB22\|BA-21\|2021-03 | NB22 | BA-21 | 23.60 | BA | BARREIRAS | 3-2021 | F | 47638.0 | 94.0 | 3735.6 | 96.7 | 98.2 |
| CHIKV\|NB23\|BA-22\|2021-06 | NB23 | BA-22 | 24.20 | BA | JACOBINA | 6-2021 | M | 19873.0 | 94.4 | 1451.0 | 96.8 | 98.3 |
| CHIKV\|NB24\|BA-23\|2021-06 | NB24 | BA-23 | 24.30 | BA | DIAS D"AVILA | 6-2021 | F | 123707.0 | 93.8 | 9189.9 | 96.7 | 98.2 |
| CHIKV\|NB25\|BA-24\|2021-04 | NB25 | BA-24 | 24.50 | BA | JOAO DOURADO | 4-2021 | F | 71258.0 | 93.4 | 4504.9 | 96.7 | 98.2 |
| CHIKV\|NB26\|BA-25\|2021-04 | NB26 | BA-25 | 24.60 | BA | JUSSARA | 4-2021 | F | 148156.0 | 95.0 | 8193.5 | 96.5 | 98.2 |
| CHIKV\|NB27\|BA-26\|2021-07 | NB27 | BA-26 | 24.80 | BA | POJUCA | 7-2021 | M | 48203.0 | 94.0 | 3785.6 | 96.7 | 98.3 |
| CHIKV\|NB28\|BA-27\|2021-06 | NB28 | BA-27 | 24.80 | BA | POJUCA | 6-2021 | F | 57357.0 | 93.9 | 3934.6 | 96.7 | 98.3 |
| CHIKV\|NB29\|BA-28\|2021-04 | NB29 | BA-28 | 25.30 | BA | PRESIDENTE DUTRA | 4-2021 | M | 71019.0 | 94.1 | 5176.7 | 96.8 | 98.3 |
| CHIKV\|NB30\|BA-29\|2021-01 | NB30 | BA-29 | 25.40 | BA | CENTRAL | 1-2021 | F | 66279.0 | 94.6 | 4807.5 | 96.7 | 98.2 |
| CHIKV\|NB31\|BA-30\|2021-07 | NB31 | BA-30 | 25.50 | BA | POJUCA | 7-2021 | M | 105284.0 | 94.2 | 6852.0 | 96.7 | 98.3 |
| CHIKV\|NB32\|BA-31\|2021-09 | NB32 | BA-31 | 26.10 | BA | POJUCA | 9-2021 | F | 92431.0 | 94.1 | 6306.1 | 96.7 | 98.2 |

| | | | | | | | | | | | | |
|---|---|---|---|---|---|---|---|---|---|---|---|---|
| CHIKV\|NB33\|BA-32\|2021-04 | NB33 | BA-32 | 26.40 | BA | JOAO DOURADO | 4-2021 | M | 115503.0 | 93.0 | 8249.5 | 96.6 | 98.1 |
| CHIKV\|NB34\|BA-33\|2021-05 | NB34 | BA-33 | 26.60 | BA | JUSSARA | 5-2021 | M | 50837.0 | 94.0 | 3464.2 | 96.6 | 98.0 |
| CHIKV\|NB35\|BA-34\|2021-06 | NB35 | BA-34 | 27.30 | BA | CAFARNAUM | 6-2021 | F | 16176.0 | 92.8 | 1076.4 | 96.8 | 98.2 |
| CHIKV\|NB36\|BA-35\|2021-06 | NB36 | BA-35 | 28.70 | BA | JACOBINA | 6-2021 | M | 35263.0 | 94.1 | 2821.5 | 96.7 | 98.3 |
| CHIKV\|BC57\|GO057\|2021-11 | BC57 | GO057 | 21.00 | GO | APARECIDA DE GOIANIA | 11-2021 | F | 66676.0 | 94.0 | 4996.5 | 96.8 | 98.2 |
| CHIKV\|BC58\|GO058\|2022-01 | BC58 | GO058 | 19.00 | GO | GOIANIA | 1-2022 | F | 9390.0 | 94.4 | 847.3 | 96.7 | 98.3 |
| CHIKV\|BC59\|GO059\|2021-12 | BC59 | GO059 | 28.00 | GO | APARECIDA DE GOIANIA | 12-2021 | F | 56752.0 | 94.3 | 3701.0 | 96.7 | 98.2 |
| CHIKV\|BC60\|GO060\|2021-12 | BC60 | GO060 | 16.00 | GO | TRINDADE | 12-2021 | F | 8140.0 | 94.4 | 712.4 | 96.7 | 98.2 |
| CHIKV\|BC61\|GO061\|2021-12 | BC61 | GO061 | 27.00 | GO | CAMPOS BELOS | 12-2021 | M | 87432.0 | 94.2 | 5213.7 | 96.7 | 98.1 |
| CHIKV\|BC62\|GO062\|2021-12 | BC62 | GO062 | 19.00 | GO | APARECIDA DE GOIANIA | 12-2021 | F | 16831.0 | 94.3 | 1210.7 | 96.7 | 98.2 |
| CHIKV\|BC63\|GO063\|2021-12 | BC63 | GO063 | 21.00 | GO | GOIANIA | 12-2021 | M | 116669.0 | 94.7 | 6980.6 | 96.6 | 98.1 |
| CHIKV\|BC64\|GO064\|2021-12 | BC64 | GO064 | 25.00 | GO | SENADOR CANEDO | 12-2021 | F | 85084.0 | 94.5 | 5579.4 | 96.6 | 98.2 |
| CHIKV\|BC65\|GO065\|2021-12 | BC65 | GO065 | 30.00 | GO | LUZIANIA | 12-2021 | F | 88300.0 | 94.4 | 5617.1 | 96.8 | 98.3 |
| CHIKV\|BC66\|GO066\|2021-12 | BC66 | GO066 | 23.00 | GO | LUZIANIA | 12-2021 | F | 71421.0 | 94.5 | 4738.0 | 96.7 | 98.2 |
| CHIKV\|BC67\|GO067\|2021-12 | BC67 | GO067 | 16.00 | GO | LUZIANIA | 12-2021 | F | 26270.0 | 94.0 | 2688.4 | 96.8 | 98.3 |
| CHIKV\|BC68\|GO068\|2021-12 | BC68 | GO068 | 25.00 | GO | LUZIANIA | 12-2021 | F | 77257.0 | 94.2 | 5221.2 | 96.8 | 98.3 |
| CHIKV\|BC69\|GO069\|2021-12 | BC69 | GO069 | 25.00 | GO | LUZIANIA | 12-2021 | M | 94412.0 | 94.4 | 6117.0 | 96.7 | 98.2 |
| CHIKV\|BC70\|GO070\|2021-12 | BC70 | GO070 | 28.00 | GO | LUZIANIA | 12-2021 | M | 69718.0 | 94.8 | 3954.7 | 96.6 | 98.3 |
| CHIKV\|BC71\|GO071\|2021-12 | BC71 | GO071 | 24.00 | GO | LUZIANIA | 12-2021 | F | 59435.0 | 94.4 | 4089.1 | 96.7 | 98.2 |
| CHIKV\|BC72\|GO072\|2021-12 | BC72 | GO072 | 20.00 | GO | VALPARAISO DE GOIAS | 12-2021 | F | 7384.0 | 94.3 | 609.2 | 96.8 | 98.3 |
| CHIKV\|BC73\|GO073\|2021-12 | BC73 | GO073 | 18.00 | GO | LUZIANIA | 12-2021 | M | 9418.0 | 93.8 | 842.8 | 96.8 | 98.3 |
| CHIKV\|BC74\|GO074\|2021-12 | BC74 | GO074 | 22.00 | GO | LUZIANIA | 12-2021 | F | 75784.0 | 94.7 | 5741.0 | 96.6 | 98.2 |
| CHIKV\|BC75\|GO075\|2021-12 | BC75 | GO075 | 26.00 | GO | LUZIANIA | 12-2021 | M | 31300.0 | 93.0 | 1737.9 | 96.7 | 98.1 |
| CHIKV\|BC76\|GO076\|2022-01 | BC76 | GO076 | 18.00 | GO | APARECIDA DE GOIANIA | 1-2022 | F | 13157.0 | 94.4 | 1106.1 | 96.8 | 98.2 |
| CHIKV\|BC77\|GO077\|2022-01 | BC77 | GO077 | 26.00 | GO | LUZIANIA | 1-2022 | F | 81286.0 | 94.1 | 5318.7 | 96.7 | 98.3 |
| CHIKV\|BC78\|GO078\|2022-01 | BC78 | GO078 | 18.00 | GO | LUZIANIA | 1-2022 | M | 69981.0 | 94.5 | 6072.8 | 96.7 | 98.2 |
| CHIKV\|BC79\|GO079\|2022-01 | BC79 | GO079 | 11.00 | GO | APARECIDA DE GOIANIA | 1-2022 | F | 3377.0 | 94.3 | 344.5 | 96.8 | 98.3 |
| CHIKV\|BC80\|GO080\|2022-01 | BC80 | GO080 | 21.00 | GO | LUZIANIA | 1-2022 | F | 2989.0 | 94.7 | 233.5 | 96.6 | 98.1 |
| CHIKV\|BC81\|GO081\|2022-01 | BC81 | GO081 | 25.00 | GO | LUZIANIA | 1-2022 | F | 91383.0 | 94.9 | 6165.5 | 96.5 | 98.2 |
| CHIKV\|BC82\|GO082\|2022-01 | BC82 | GO082 | 17.00 | GO | CAMPOS BELOS | 1-2022 | F | 48503.0 | 94.5 | 4618.1 | 96.7 | 98.1 |
| CHIKV\|BC83\|GO083\|2022-01 | BC83 | GO083 | 23.00 | GO | GOIANIA | 1-2022 | F | 116617.0 | 94.0 | 8253.4 | 96.8 | 98.2 |
| CHIKV\|BC84\|GO084\|2022-01 | BC84 | GO084 | 25.00 | GO | LUZIANIA | 1-2022 | M | 91278.0 | 94.1 | 6363.5 | 96.7 | 98.3 |
| CHIKV\|BC85\|GO085\|2021-05 | BC85 | GO085 | 21.00 | GO | PEROLANDIA | 5-2021 | F | 96788.0 | 94.6 | 6721.9 | 96.6 | 98.2 |

| | | | | | | | | | | | | |
|---|---|---|---|---|---|---|---|---|---|---|---|---|
| CHIKV\|BC86\|GO086\|2021-05 | BC86 | GO086 | 25.00 | GO | PEROLANDIA | 5-2021 | M | 109908.0 | 93.9 | 7608.2 | 96.8 | 98.3 |
| CHIKV\|BC87\|GO087\|2021-05 | BC87 | GO087 | 26.00 | GO | BOM JESUS DE GOIAS | 5-2021 | F | 65882.0 | 94.5 | 4546.6 | 96.8 | 98.2 |
| CHIKV\|BC88\|GO088\|2021-05 | BC88 | GO088 | 28.00 | GO | BOM JESUS DE GOIAS | 5-2021 | F | 19663.0 | 94.4 | 1358.4 | 96.7 | 98.2 |
| CHIKV\|BC89\|GO089\|2021-06 | BC89 | GO089 | 18.00 | GO | ANAPOLIS | 6-2021 | F | 27680.0 | 94.1 | 2883.4 | 96.7 | 98.3 |
| CHIKV\|BC90\|GO090\|2021-09 | BC90 | GO090 | 28.00 | GO | APARECIDA DE GOIANIA | 9-2021 | F | 70631.0 | 94.1 | 4804.8 | 96.7 | 98.2 |
| CHIKV\|BC91\|GO091\|2021-11 | BC91 | GO091 | 21.00 | GO | APARECIDA DE GOIANIA | 11-2021 | F | 96503.0 | 94.3 | 6692.1 | 96.6 | 98.2 |
| CHIKV\|BC92\|GO092\|2021-11 | BC92 | GO092 | 24.00 | GO | APARECIDA DE GOIANIA | 11-2021 | M | 84959.0 | 94.0 | 6206.1 | 96.7 | 98.3 |
| CHIKV\|BC93\|GO093\|2021-11 | BC93 | GO093 | 24.00 | GO | GOIANIA | 11-2021 | F | 18253.0 | 94.2 | 1313.6 | 96.8 | 98.2 |
| CHIKV\|BC94\|GO094\|2021-11 | BC94 | GO094 | 23.00 | GO | GOIANIA | 11-2021 | M | 79545.0 | 94.2 | 5825.6 | 96.8 | 98.2 |
| CHIKV\|BC95\|GO095\|2022-01 | BC95 | GO095 | 22.00 | GO | TRINDADE | 1-2022 | F | 29295.0 | 94.3 | 1791.4 | 96.8 | 98.2 |
| CHIKV\|BC96\|GO096\|2021-12 | BC96 | GO096 | 22.00 | GO | GOIANIA | 12-2021 | M | 725.0 | 93.9 | 58.1 | 96.8 | 98.3 |
| CHIKV\|MA\|BC01\|2022-06 | BC01 | - | 22.00 | MA | SAO LUIS | 6-2022 | F | 15380 | 92.6 | 964.7 | 96.6 | 98.1 |
| CHIKV\|MA\|BC02\|2022-06 | BC02 | - | 24.00 | MA | SAO LUIS | 6-2022 | F | 15713 | 88.8 | 1061.1 | 96.7 | 98 |
| CHIKV\|MA\|BC03\|2022-06 | BC03 | - | 23.00 | MA | SAO LUIS | 6-2022 | M | 9121 | 90.9 | 590 | 96.8 | 98.2 |
| CHIKV\|MA\|BC04\|2022-06 | BC04 | - | 26.00 | MA | SAO LUIS | 6-2022 | F | 17624 | 82.8 | 1306 | 96.7 | 98.1 |
| CHIKV\|MA\|BC05\|2022-05 | BC05 | - | 24.00 | MA | SAO LUIS | 5-2022 | M | 16780 | 87.8 | 1125.5 | 96.8 | 98.1 |
| CHIKV\|MA\|BC19\|2022-06 | BC19 | - | 22.00 | MA | BACABAL | 6-2022 | F | 24423 | 90.7 | 1658.1 | 96.7 | 98.3 |
| CHIKV\|MA\|BC20\|2022-06 | BC20 | - | 22.00 | MA | CAXIAS | 6-2022 | F | 26619 | 87.8 | 1867.7 | 96.5 | 97.8 |
| CHIKV\|MA\|BC21\|2022-06 | BC21 | - | 23.00 | MA | CAXIAS | 6-2022 | M | 18226 | 86.9 | 1172.8 | 96.5 | 97.8 |
| CHIKV\|MA\|BC22\|2022-06 | BC22 | - | 25.00 | MA | SAO LUIS | 6-2022 | F | 16062 | 84.7 | 1134.6 | 96.7 | 98 |
| CHIKV\|MA\|BC23\|2022-05 | BC23 | - | 24.00 | MA | SAO LUIS | 5-2022 | M | 18760 | 88.5 | 1365.7 | 96.7 | 98 |
| CHIKV\|MA\|BC24\|2022-05 | BC24 | - | 24.00 | MA | SAO LUIS | 5-2022 | F | 9620 | 81.2 | 781.5 | 96.6 | 98.1 |
| CHIKV\|MA\|BC25\|2022-06 | BC25 | - | 24.00 | MA | SAO LUIS | 6-2022 | M | 17308 | 87.4 | 1304.8 | 96.7 | 98.1 |
| CHIKV\|MA\|BC26\|2022-06 | BC26 | - | 23.00 | MA | SAO LUIS | 6-2022 | F | 24030 | 93.2 | 1545.8 | 96.7 | 98.2 |
| CHIKV\|MA\|BC40\|2022-06 | BC40 | - | 25.00 | MA | SAO LUIS | 6-2022 | F | 17687 | 85.6 | 1227.8 | 96.7 | 98.1 |
| CHIKV\|MA\|BC41\|2022-06 | BC41 | - | 24.00 | MA | SAO LUIS | 6-2022 | F | 31300 | 87.6 | 1940.4 | 96.7 | 98.1 |
| CHIKV\|MA\|BC42\|2022-06 | BC42 | - | 22.00 | MA (SP) | CAJAMAR | 6-2022 | M | 27099 | 86.5 | 1959.9 | 96.8 | 98.3 |
| CHIKV\|MA\|BC43\|2022-06 | BC43 | - | 25.00 | MA | SAO LUIS | 6-2022 | M | 33729 | 85.3 | 2364.5 | 96.7 | 98.1 |
| CHIKV\|MA\|BC44\|2022-06 | BC44 | - | 24.00 | MA | SANTA INES | 6-2022 | F | 18897 | 85.5 | 1250.6 | 96.5 | 98.1 |
| CHIKV\|MA\|BC49\|2022-05 | BC49 | - | 22.00 | MA | SAO LUIS | 5-2022 | F | 17094 | 89.4 | 1078.1 | 96.7 | 98.1 |
| CHIKV\|MA\|BC50\|2022-05 | BC50 | - | 21.00 | MA | TIMON | 5-2022 | M | 12600 | 91.5 | 821 | 96.8 | 98.2 |
| CHIKV\|MA\|BC51\|2022-06 | BC51 | - | 24.00 | MA | SAO LUIS | 6-2022 | M | 11100 | 83 | 756.4 | 96.7 | 98.1 |
| CHIKV\|MA\|BC52\|2022-05 | BC52 | - | 21.00 | MA (MG) | SANTA JULIANA | 5-2022 | M | 12248 | 90.2 | 822.3 | 96.6 | 98 |

| | | | | | | | | | | | | |
|---|---|---|---|---|---|---|---|---|---|---|---|---|
| CHIKV\|MA\|BC53\|2022-05 | BC53 | - | 24.00 | MA | SAO LUIS | 5-2022 | M | 20722 | 85.5 | 1336.9 | 96.5 | 97.9 |
| CHIKV\|MA\|BC54\|2022-05 | BC54 | - | 27.00 | MA | SAO LUIS | 5-2022 | M | 9404 | 84.8 | 664.6 | 96.4 | 97.8 |
| CHIKV\|MA\|BC55\|2022-05 | BC55 | - | 21.00 | MA | SAO LUIS | 5-2022 | M | 14551 | 92.6 | 944.8 | 96.8 | 98.3 |
| CHIKV\|MA\|BC56\|2022-05 | BC56 | - | 24.00 | MA | SAO LUIS | 5-2022 | F | 13439 | 87.4 | 955.4 | 96.7 | 98 |
| CHIKV\|MA\|BC57\|2022-05 | BC57 | - | 24.00 | MA | SAO LUIS | 5-2022 | F | 12884 | 86.8 | 862.1 | 96.8 | 98.1 |
| CHIKV\|MA\|BC58\|2022-05 | BC58 | - | 26.00 | MA (RR) | BOA VISTA | 5-2022 | F | 16879 | 88.3 | 1117.1 | 96.5 | 98.1 |
| CHIKV\|MA\|BC59\|2022-05 | BC59 | - | 23.00 | MA (PA) | BELEM | 5-2022 | M | 13390 | 87.3 | 885.8 | 96.7 | 98.1 |
| CHIKV\|MA\|BC69\|2022-05 | BC69 | - | 23.00 | MA | SAO LUIS | 5-2022 | M | 13059 | 89.8 | 846.9 | 96.7 | 98.2 |
| CHIKV\|MA\|BC70\|2022-05 | BC70 | - | 26.00 | MA | SAO LUIS | 5-2022 | F | 9868 | 85.1 | 664.1 | 96.7 | 98 |
| CHIKV\|MA\|BC71\|2022-05 | BC71 | - | 25.00 | MA | SAO LUIS | 5-2022 | F | 13689 | 88 | 898.7 | 96.6 | 98 |
| CHIKV\|MA\|BC72\|2022-05 | BC72 | - | 20.00 | MA | SAO LUIS | 5-2022 | M | 11537 | 92.6 | 766.9 | 96.8 | 98.2 |
| CHIKV\|MA\|BC73\|2022-05 | BC73 | - | 20.00 | MA | SAO LUIS | 5-2022 | F | 15871 | 93.1 | 1042.7 | 96.8 | 98.3 |
| CHIKV\|MA\|BC74\|2022-05 | BC74 | - | 27.00 | MA | SAO LUIS | 5-2022 | F | 17914 | 77.7 | 1472.4 | 96.6 | 97.9 |
| CHIKV\|MA\|BC75\|2022-05 | BC75 | - | 25.00 | MA | SAO LUIS | 5-2022 | M | 18346 | 85.7 | 1212.6 | 96.6 | 97.9 |
| CHIKV\|MA\|BC76\|2022-05 | BC76 | - | 23.00 | MA | SAO LUIS | 5-2022 | F | 31268 | 84.7 | 2207.1 | 96.5 | 97.9 |
| CHIKV\|MA\|BC89\|2022-05 | BC89 | - | 20.00 | MA | SAO LUIS | 5-2022 | M | 22535 | 92.8 | 1488.8 | 96.7 | 98.1 |
| CHIKV\|MS\|BC11\|2022-04 | BC11 | - | 18.00 | MS | CAMPO GRANDE | 4-2022 | F | 27643 | 94.1 | 1657 | - | - |
| CHIKV\|MS\|BC12\|2022-04 | BC12 | - | 27.00 | MS | CAMPO GRANDE | 4-2022 | M | 48153 | 94.3 | 2699.4 | - | - |
| CHIKV\|PB\|PB017\|NBC17\|2021-05 | NBC17 | PB017 | 27.00 | PB | REMIGIO | 5-2021 | M | 172377 | 94.2 | 9450.2 | 96.9 | 98.4 |
| CHIKV\|PB\|PB018\|NBC18\|2021-06 | NBC18 | PB018 | 21.00 | PB | CAPIM | 6-2021 | F | 108135 | 94.1 | 6318 | 96.9 | 98.3 |
| CHIKV\|PB\|PB019\|NBC19\|2021-07 | NBC19 | PB019 | 18.00 | PB | JOAO PESSOA | 7-2021 | M | ~1600 | 91.6 | 170 | 96.8 | 98.3 |
| CHIKV\|PB\|PB020\|NBC20\|2021-06 | NBC20 | PB020 | 16.00 | PB | JOAO PESSOA | 6-2021 | M | 57899 | 94 | 4989.4 | 96.9 | 98.4 |
| CHIKV\|PB\|PB021\|NBC21\|2021-06 | NBC21 | PB021 | 20.00 | PB | JOAO PESSOA | 6-2021 | F | 65560 | 94 | 4074.3 | 96.8 | 98.4 |
| CHIKV\|PB\|PB022\|NBC22\|2021-05 | NBC22 | PB022 | 18.00 | PB | JOAO PESSOA | 5-2021 | F | 23352 | 93.9 | 1688.4 | 96.8 | 98.4 |
| CHIKV\|PB\|PB023\|NBC23\|2021-06 | NBC23 | PB023 | 28.00 | PB | JOAO PESSOA | 6-2021 | M | 138786 | 94.1 | 7473.3 | 96.9 | 98.4 |
| CHIKV\|PB\|PB024\|NBC24\|2021-06 | NBC24 | PB024 | 18.00 | PB | SUME | 6-2021 | M | 69942 | 94.3 | 5616.9 | 96.6 | 98.2 |
| CHIKV\|PB\|PB025\|NBC25\|2021-05 | NBC25 | PB025 | 15.00 | PB | CONDE | 5-2021 | F | 53626 | 94.3 | 4135.1 | 96.8 | 98.3 |
| CHIKV\|PB\|PB026\|NBC26\|2021-06 | NBC26 | PB026 | 20.00 | PB | CAAPORA | 6-2021 | F | 109771 | 94.5 | 6857.6 | 96.8 | 98.3 |
| CHIKV\|PB\|PB027\|NBC27\|2021-06 | NBC27 | PB027 | 17.00 | PB | JOAO PESSOA | 6-2021 | F | 61673 | 94 | 4704 | 96.8 | 98.4 |
| CHIKV\|PB\|PB028\|NBC28\|2021-07 | NBC28 | PB028 | 22.00 | PB | LUCENA | 7-2021 | F | 51416 | 94.3 | 3498.7 | 96.8 | 98.2 |
| CHIKV\|PB\|PB029\|NBC29\|2021-08 | NBC29 | PB029 | 14.00 | PB | JOAO PESSOA | 8-2021 | M | 10559 | 93.8 | 911.9 | 96.8 | 98.3 |
| CHIKV\|SP\|PB030\|NBC30\|2021-07 | NBC30 | PB030 | 23.00 | PB (SP) | SAO PAULO | 7-2021 | M | 93533 | 93.8 | 5672.5 | 96.8 | 98.4 |

| | | | | | | | | | | | | |
|---|---|---|---|---|---|---|---|---|---|---|---|---|
| CHIKV\|PB\|PB031\|NBC31\|2021-08 | NBC31 | PB031 | 25.00 | PB | JOAO PESSOA | 8-2021 | M | 87700 | 93.9 | 5246.2 | 96.7 | 98.3 |
| CHIKV\|PB\|PB032\|NBC32\|2021-08 | NBC32 | PB032 | 18.00 | PB | CONDE | 8-2021 | M | 53784 | 93.9 | 3875.3 | 96.8 | 98.4 |
| CHIKV\|PB\|PB033\|NBC33\|2021-08 | NBC33 | PB033 | 13.00 | PB | ALAGOINHA | 8-2021 | M | ~1300 | 91.6 | 160 | 96.8 | 98.2 |
| CHIKV\|PB\|PB034\|NBC34\|2021-09 | NBC34 | PB034 | 22.00 | PB | SANTA RITA | 9-2021 | M | 95964 | 93.9 | 5833.5 | 96.7 | 98.4 |
| CHIKV\|PB\|PB035\|NBC35\|2021-09 | NBC35 | PB035 | 18.00 | PB | JOAO PESSOA | 9-2021 | M | 22917 | 93.9 | 1625.7 | 96.8 | 98.4 |
| CHIKV\|PB\|PB036\|NBC36\|2021-09 | NBC36 | PB036 | 18.00 | PB | JOAO PESSOA | 9-2021 | F | 39209 | 94.3 | 3157.3 | 96.8 | 98.2 |
| CHIKV\|PB\|PB037\|NBC37\|2021-08 | NBC37 | PB037 | 20.00 | PB | JOAO PESSOA | 8-2021 | F | 54302 | 94.1 | 3620.1 | 96.9 | 98.3 |
| CHIKV\|PB\|PB038\|NBC38\|2021-09 | NBC38 | PB038 | 18.00 | PB | CAAPORA | 9-2021 | M | 62611 | 94.2 | 4394 | 96.9 | 98.4 |
| CHIKV\|PB\|PB039\|NBC39\|2021-10 | NBC39 | PB039 | 21.00 | PB | ALAGOA GRANDE | 10-2021 | F | 1361 | 93.7 | 89.9 | 97 | 98.4 |
| CHIKV\|PE\|5572\|NBC01\|2021-10 | NBC01 | 5572 | 27.62 | PE | BARREIROS | 10-2021 | F | ~2000 | 89 | 200 | 96.7 | 98.1 |
| CHIKV\|PE\|5542\|NBC02\|2021-10 | NBC02 | 5542 | 28.34 | PE | IPOJUCA | 10-2021 | M | 46980 | 94.2 | 3559.7 | 96.9 | 98.4 |
| CHIKV\|PE\|4240\|NBC03\|2021-08 | NBC03 | 4240 | 27.45 | PE | BARREIROS | 8-2021 | M | ~1700 | 91.1 | 180 | 96.8 | 98.1 |
| CHIKV\|PE\|4258\|NBC04\|2021-08 | NBC04 | 4258 | 24.87 | PE | GOIANIA | 8-2021 | M | 99774 | 93.9 | 6987.5 | 97 | 98.5 |
| CHIKV\|PE\|5439\|NBC05\|2021-10 | NBC05 | 5439 | 27.87 | PE | BARREIROS | 10-2021 | M | 68108 | 93.8 | 4425.5 | 96.8 | 98.3 |
| CHIKV\|PE\|5462\|NBC06\|2021-10 | NBC06 | 5462 | 25.05 | PE | SOURE | 10-2021 | F | 64105 | 94.3 | 5166.8 | 96.8 | 98.3 |
| CHIKV\|PE\|5464\|NBC07\|2021-10 | NBC07 | 5464 | 22.08 | PE | FERNANDO DE NORONHA | 10-2021 | F | 18131 | 93.8 | 1739.8 | 96.8 | 98.3 |
| CHIKV\|PE\|5313\|NBC08\|2021-10 | NBC08 | 5313 | 26.46 | PE | VICENCIA | 10-2021 | M | 53674 | 93.9 | 3387.5 | 96.9 | 98.4 |
| CHIKV\|PE\|5342\|NBC09\|2021-09 | NBC09 | 5342 | 27.31 | PE | CONDADO | 9-2021 | F | 979 | 89.4 | 70.3 | 97.1 | 98.5 |
| CHIKV\|PE\|5348\|NBC10\|2021-09 | NBC10 | 5348 | 24.32 | PE | FERNANDO DE NORONHA | 9-2021 | M | 16364 | 93.8 | 1306.4 | 96.9 | 98.4 |
| CHIKV\|PE\|5375\|NBC11\|2021-10 | NBC11 | 5375 | 25.04 | PE | AGUA PRETA | 10-2021 | F | 50206 | 93.9 | 3518.3 | 96.7 | 98.2 |
| CHIKV\|PE\|5118\|NBC12\|2021-09 | NBC12 | 5118 | 26.31 | PE | PAULISTA | 9-2021 | F | ~400 | 91.3 | 200 | 96.7 | 98.3 |
| CHIKV\|PE\|5126\|NBC13\|2021-09 | NBC13 | 5126 | 26.31 | PE | BARREIROS | 9-2021 | F | 43294 | 94.9 | 3159.1 | 96.7 | 98.1 |
| CHIKV\|PE\|5127\|NBC14\|2021-09 | NBC14 | 5127 | 26.50 | PE | BARREIROS | 9-2021 | M | 39295 | 93.8 | 2557.8 | 96.8 | 98.3 |
| CHIKV\|PE\|4641\|NBC15\|2021-08 | NBC15 | 4641 | 25.11 | PE | JABOATAO DOS GUARARAPES | 8-2021 | M | 52875 | 94.1 | 3948.2 | 96.9 | 98.3 |
| CHIKV\|PE\|4664\|NBC16\|2021-08 | NBC16 | 4664 | 23.02 | PE | FEIRA NOVA | 8-2021 | F | 38012 | 94.4 | 2653.8 | 96.8 | 98.2 |
| CHIKV\|PI06\|NBC06\|2021-07 | NB06 | PI06 | 30.00 | PI | TERESINA | 7-2021 | M | 1779643 | 93.4 | 129706.9 | 96.6 | 98.1 |
| CHIKV\|PI07\|NBC07\|2021-08 | NB07 | PI07 | 30.00 | PI | TERESINA | 8-2021 | M | 1681440 | 90.8 | 103651.5 | 96.7 | 98 |
| CHIKV\|PI\|NB01\|2022-02 | NB01 | - | 25.00 | PI | TERESINA | 2-2022 | F | 12201 | 92.6 | 786.7 | 96.6 | 97.9 |
| CHIKV\|PI\|NB02\|2022-01 | NB02 | - | 26.00 | PI | TERESINA | 1-2022 | F | 20517 | 94.5 | 1583.2 | 96.5 | 98.1 |
| CHIKV\|PI\|NB03\|2022-02 | NB03 | - | 27.00 | PI | TERESINA | 2-2022 | F | 10368 | 94.6 | 718.5 | 96.6 | 98.2 |
| CHIKV\|PI\|NB04\|2022-02 | NB04 | - | 28.00 | PI | TERESINA | 2-2022 | F | 16232 | 94 | 1110 | 96.6 | 98.2 |
| CHIKV\|PI\|NB06\|2022-03 | NB06 | - | 26.00 | PI | TERESINA | 3-2022 | M | 19842 | 94.1 | 1492.3 | 96.6 | 98.1 |

| | | | | | | | | | | | | |
|---|---|---|---|---|---|---|---|---|---|---|---|---|
| CHIKV\|PI\|NB07\|2022-03 | NB07 | - | 29.00 | PI | TERESINA | 3-2022 | M | 10088 | 94.2 | 649.1 | 96.6 | 98.2 |
| CHIKV\|PI\|NB08\|2022-03 | NB08 | - | 27.00 | PI | TERESINA | 3-2022 | F | 16292 | 94.3 | 1114.2 | 96.6 | 98.1 |
| CHIKV\|PI\|NB09\|2022-03 | NB09 | - | 29.00 | PI | TERESINA | 3-2022 | F | 13176 | 94 | 1051.1 | 96.6 | 98.2 |
| CHIKV\|PI\|NB10\|2022-03 | NB10 | - | 27.00 | PI | TERESINA | 3-2022 | F | 21506 | 94.8 | 1489.1 | 96.7 | 98.2 |
| CHIKV\|PI\|NB11\|2022-03 | NB11 | - | 29.00 | PI | TERESINA | 3-2022 | M | 13639 | 94 | 1087 | 96.6 | 98.1 |
| CHIKV\|PI\|NB12\|2022-03 | NB12 | - | 27.00 | PI | TERESINA | 3-2022 | M | 18907 | 94 | 1496.1 | 96.6 | 98.2 |
| CHIKV\|PI\|NB13\|2022-03 | NB13 | - | 27.00 | PI | TERESINA | 3-2022 | F | 18498 | 95.3 | 1415.6 | 96.3 | 98 |
| CHIKV\|PI\|NB14\|2022-03 | NB14 | - | 29.00 | PI | TERESINA | 3-2022 | F | 12553 | 93.8 | 894.8 | 96.6 | 98.2 |
| CHIKV\|PI\|NB15\|2022-03 | NB15 | - | 28.00 | PI | TERESINA | 3-2022 | F | 10043 | 94.5 | 739.8 | 96.6 | 98.1 |
| CHIKV\|PI\|NB16\|2022-03 | NB16 | - | 20.00 | PI | TERESINA | 3-2022 | F | 13719 | 78.4 | 805.1 | 95.7 | 96.9 |
| CHIKV\|PI\|NB17\|2022-03 | NB17 | - | 29.00 | PI | TERESINA | 3-2022 | F | 15330 | 94.1 | 1093.2 | 96.6 | 98.2 |
| CHIKV\|PI\|NB18\|2022-03 | NB18 | - | 27.00 | PI | TERESINA | 3-2022 | M | 12501 | 94.2 | 1060.9 | 96.6 | 98.1 |
| CHIKV\|PI\|NB19\|2022-03 | NB19 | - | 30.00 | PI | TERESINA | 3-2022 | F | 19575 | 94.2 | 1472.8 | 96.7 | 98.2 |
| CHIKV\|PI\|NB20\|2022-03 | NB20 | - | 30.00 | PI | TERESINA | 3-2022 | F | 21103 | 94.9 | 1559.4 | 96.5 | 97.9 |
| CHIKV\|PI\|NB21\|2022-03 | NB21 | - | 29.00 | PI | TERESINA | 3-2022 | F | 22248 | 94.5 | 1511.5 | 96.5 | 98.1 |
| CHIKV\|PI\|NB22\|2022-03 | NB22 | - | 28.00 | PI | TERESINA | 3-2022 | M | 18295 | 93.9 | 1365.1 | 96.8 | 98.3 |
| CHIKV\|PI\|NB23\|2022-03 | NB23 | - | 28.00 | PI | TERESINA | 3-2022 | M | 5089 | 94.3 | 373.8 | 96.7 | 98.3 |
| CHIKV\|PI\|NB24\|2022-03 | NB24 | - | 30.00 | PI | TERESINA | 3-2022 | F | 8309 | 93.6 | 552.9 | 96.4 | 98.1 |
| CHIKV\|PI\|NB25\|2022-03 | NB25 | - | 28.00 | PI | TERESINA | 3-2022 | F | 9121 | 93.1 | 660.9 | 96.6 | 98.1 |
| CHIKV\|PI\|NB26\|2022-03 | NB26 | - | 28.00 | PI | TERESINA | 3-2022 | M | 14851 | 94.9 | 1024.1 | 96.4 | 98.1 |
| CHIKV\|PI\|NB27\|2022-03 | NB27 | - | 26.00 | PI | UNIÃO | 3-2022 | F | 15621 | 93.9 | 1065.4 | 96.6 | 98.1 |
| CHIKV\|PI\|NB28\|2022-03 | NB28 | - | 28.00 | PI | TERESINA | 3-2022 | F | 7590 | 94 | 563.8 | 96.6 | 98.2 |
| CHIKV\|PI\|NB29\|2022-04 | NB29 | - | 29.00 | PI | TERESINA | 4-2022 | M | 11405 | 94 | 835.2 | 96.6 | 98.2 |
| CHIKV\|PI\|NB30\|2022-04 | NB30 | - | 28.00 | PI | TERESINA | 4-2022 | F | 16892 | 93.3 | 1133 | 96.6 | 98.2 |
| CHIKV\|PI\|NB31\|2022-04 | NB31 | - | 28.00 | PI | TERESINA | 4-2022 | M | 12862 | 94.1 | 901.1 | 96.6 | 98.2 |
| CHIKV\|PI\|NB32\|2022-04 | NB32 | - | 26.00 | PI | TERESINA | 4-2022 | M | 16092 | 94.1 | 1224.4 | 96.6 | 98.2 |
| CHIKV\|PI\|NB33\|2022-03 | NB33 | - | 30.00 | PI | TERESINA | 3-2022 | F | 9678 | 93.3 | 690.7 | 96.6 | 98.1 |
| CHIKV\|PI\|NB34\|2022-04 | NB34 | - | 29.00 | PI | TERESINA | 4-2022 | F | 5582 | 93.1 | 500.2 | 96.6 | 98.1 |
| CHIKV\|PI\|NB35\|2022-03 | NB35 | - | 25.00 | PI | TERESINA | 3-2022 | M | 8226 | 92.5 | 622.7 | 96.7 | 98.2 |
| CHIKV\|PI\|NB37\|2022-03 | NB37 | - | 27.00 | PI | TERESINA | 3-2022 | F | 7864 | 76.2 | 575.1 | 96.4 | 97.7 |
| CHIKV\|PI\|NB38\|2022-04 | NB38 | - | 28.00 | PI | TERESINA | 4-2022 | F | 8909 | 94.3 | 623.9 | 96.6 | 98.1 |
| CHIKV\|PI\|NB39\|2022-03 | NB39 | - | 29.00 | PI | TERESINA | 3-2022 | F | 6820 | 94.9 | 502.9 | 96.5 | 98 |
| CHIKV\|PI\|NB40\|2022-04 | NB40 | - | 30.00 | PI | TERESINA | 4-2022 | M | 6015 | 94.3 | 390.1 | 96.6 | 98 |

| | | | | | | | | | | | | |
|---|---|---|---|---|---|---|---|---|---|---|---|---|
| CHIKV\|PI\|NB41\|2022-04 | NB41 | - | 24.00 | PI | TERESINA | 4-2022 | F | 5361 | 94.4 | 433.2 | 96.6 | 98.1 |
| CHIKV\|PI\|NB42\|2022-04 | NB42 | - | 25.00 | PI | TERESINA | 4-2022 | F | 6416 | 94 | 542.3 | 96.6 | 98.1 |
| CHIKV\|PI\|NB43\|2022-04 | NB43 | - | 29.00 | PI | TERESINA | 4-2022 | M | 9979 | 94.3 | 658.6 | 96.6 | 98.1 |
| CHIKV\|PI\|NB44\|2022-04 | NB44 | - | 30.00 | PI | TERESINA | 4-2022 | F | 6639 | 94.2 | 420.7 | 96.6 | 98.2 |
| CHIKV\|PI\|NB45\|2022-04 | NB45 | - | 26.00 | PI | TERESINA | 4-2022 | M | 10682 | 92.8 | 697.8 | 96.6 | 98.1 |
| CHIKV\|PI\|NB46\|2022-04 | NB46 | - | 30.00 | PI | TERESINA | 4-2022 | F | 5700 | 94.4 | 418.9 | 96.6 | 98.1 |
| CHIKV\|PI\|NB47\|2022-04 | NB47 | - | 28.00 | PI | TERESINA | 4-2022 | F | 6657 | 94 | 461.9 | 96.6 | 98.2 |
| CHIKV\|PI\|NB48\|2022-04 | NB48 | - | 28.00 | PI | TERESINA | 4-2022 | M | 4820 | 93.7 | 343.9 | 96.5 | 98 |
| CHIKV\|PI\|NB49\|2022-04 | NB49 | - | 27.00 | PI | TERESINA | 4-2022 | M | 18574 | 92.6 | 1357.4 | 96.6 | 98 |
| CHIKV\|PI\|NB50\|2022-04 | NB50 | - | 28.00 | PI | TERESINA | 4-2022 | F | 15671 | 90.5 | 1070.6 | 96.6 | 98 |
| CHIKV\|PI\|NB51\|2022-04 | NB51 | - | 30.00 | PI | TERESINA | 4-2022 | M | 14221 | 82.3 | 1047.3 | 96.4 | 97.7 |
| CHIKV\|PI\|NB52\|2022-04 | NB52 | - | 28.00 | PI | TERESINA | 4-2022 | F | 26750 | 92.9 | 2036 | 96.6 | 98.2 |
| CHIKV\|PI\|NB53\|2022-04 | NB53 | - | 30.00 | PI | TERESINA | 4-2022 | F | 25594 | 90.2 | 1726 | 96.5 | 98 |
| CHIKV\|PI\|NB54\|2022-04 | NB54 | - | 27.00 | PI | TERESINA | 4-2022 | F | 15116 | 91.8 | 1156.6 | 96.3 | 97.7 |
| CHIKV\|PI\|NB55\|2022-04 | NB55 | - | 28.00 | PI | BOQUEIRAO DO PIAUI | 4-2022 | F | 18117 | 91.6 | 1205.1 | 96.4 | 97.8 |
| CHIKV\|PI\|NB56\|2022-04 | NB56 | - | 27.00 | PI | TERESINA | 4-2022 | M | 18117 | 91.6 | 1205.1 | 96.4 | 97.8 |
| CHIKV\|PI\|NB57\|2022-04 | NB57 | - | 26.00 | PI | TERESINA | 4-2022 | F | 17821 | 92.3 | 1181.9 | 96.5 | 97.9 |
| CHIKV\|PI\|NB58\|2022-04 | NB58 | - | 25.00 | PI | TERESINA | 4-2022 | F | 12749 | 91.9 | 990.4 | 96.2 | 97.7 |
| CHIKV\|PI\|NB59\|2022-04 | NB59 | - | 28.00 | PI | TERESINA | 4-2022 | M | 24323 | 86.9 | 1725.7 | 96.5 | 97.8 |
| CHIKV\|PI\|NB60\|2022-04 | NB60 | - | 28.00 | PI | TERESINA | 4-2022 | M | 20071 | 90.1 | 1482.3 | 96.5 | 97.8 |
| CHIKV\|PI\|NB61\|2022-04 | NB61 | - | 30.00 | PI | TERESINA | 4-2022 | M | 22407 | 82.2 | 1565.9 | 96.4 | 97.5 |
| CHIKV\|PI\|NB62\|2022-04 | NB62 | - | 28.00 | PI | TERESINA | 4-2022 | M | 21006 | 89.8 | 1422.3 | 96.4 | 97.6 |
| CHIKV\|PI\|NB63\|2022-04 | NB63 | - | 30.00 | PI | TERESINA | 4-2022 | F | 15311 | 90.5 | 1148.7 | 96.6 | 98.1 |
| CHIKV\|PI\|NB64\|2022-04 | NB64 | - | 26.00 | PI | TERESINA | 4-2022 | F | 16081 | 82.6 | 1143 | 96.2 | 97.7 |
| CHIKV\|PI\|NB65\|2022-04 | NB65 | - | 27.00 | PI | TERESINA | 4-2022 | M | 18838 | 90.2 | 1335.7 | 96.3 | 97.7 |
| CHIKV\|PI\|NB66\|2022-04 | NB66 | - | 29.00 | PI | TERESINA | 4-2022 | M | 13921 | 86.9 | 1080.3 | 96.5 | 97.9 |
| CHIKV\|PI\|NB67\|2022-04 | NB67 | - | 29.00 | PI | TERESINA | 4-2022 | F | 14815 | 90.6 | 1035.5 | 96.6 | 98 |
| CHIKV\|PI\|NB68\|2022-04 | NB68 | - | 25.00 | PI | TERESINA | 4-2022 | F | 16313 | 92.8 | 1283.9 | 96.6 | 98 |
| CHIKV\|PI\|NB69\|2022-04 | NB69 | - | 26.00 | PI | AGRICOLANDIA | 4-2022 | M | 12919 | 85.2 | 995.4 | 96.4 | 97.9 |
| CHIKV\|PI\|NB70\|2022-04 | NB70 | - | 30.00 | PI | TERESINA | 4-2022 | F | 22456 | 88.9 | 1575.6 | 96.6 | 97.8 |
| CHIKV\|PI\|NB71\|2022-04 | NB71 | - | 25.00 | PI | TERESINA | 4-2022 | M | 16212 | 90.3 | 1247.5 | 96.5 | 97.9 |
| CHIKV\|PI\|NB72\|2022-04 | NB72 | - | 27.00 | PI | TERESINA | 4-2022 | F | 21832 | 90.6 | 1214.2 | 96.6 | 98 |
| CHIKV\|PI\|NB73\|2022-05 | NB73 | - | 26.00 | PI | TERESINA | 5-2022 | F | 19482 | 92.2 | 1284 | 96.6 | 98.1 |

| | | | | | | | | | | | | |
|---|---|---|---|---|---|---|---|---|---|---|---|---|
| CHIKV\|PI\|NB74\|2022-04 | NB74 | - | 25.00 | PI | TERESINA | 4-2022 | F | 31171 | 86.5 | 2249.6 | 96.4 | 97.7 |
| CHIKV\|PI\|NB75\|2022-05 | NB75 | - | 28.00 | PI | TERESINA | 5-2022 | F | 24562 | 85.3 | 1733.9 | 96.4 | 97.6 |
| CHIKV\|PI\|NB76\|2022-04 | NB76 | - | 28.00 | PI | TERESINA | 4-2022 | M | 29219 | 89.5 | 2059.5 | 96.6 | 98.1 |
| CHIKV\|PI\|NB77\|2022-05 | NB77 | - | 29.00 | PI | TERESINA | 5-2022 | F | 18968 | 84.3 | 1381.1 | 96.7 | 98.1 |
| CHIKV\|PI\|NB78\|2022-05 | NB78 | - | 30.00 | PI | TERESINA | 5-2022 | F | 32919 | 90.2 | 2260.1 | 96.8 | 98.1 |
| CHIKV\|PI\|NB79\|2022-05 | NB79 | - | 16.00 | PI | TERESINA | 5-2022 | M | 4406 | 94.2 | 558.5 | 96.6 | 98.2 |
| CHIKV\|PI\|NB80\|2022-05 | NB80 | - | 27.00 | PI | TERESINA | 5-2022 | M | 22919 | 91.7 | 1810.6 | 96.6 | 98 |
| CHIKV\|PI\|NB81\|2022-06 | NB81 | - | 25.00 | PI | PARNAIBA | 6-2022 | M | 23277 | 89.3 | 1415.7 | 96.6 | 98 |
| CHIKV\|PI\|NB82\|2022-06 | NB82 | - | 28.00 | PI | ALVORADA DO GURGUEIA | 6-2022 | F | 26009 | 95.2 | 2044.5 | 96.5 | 98 |
| CHIKV\|PI\|NB84\|2022-06 | NB84 | - | 28.00 | PI (RIO GRANDE DO SUL) | ELDORADO DO SUL | 6-2022 | F | 27053 | 88.1 | 1814.4 | 96.5 | 98 |
| CHIKV\|PI\|NB85\|2022-06 | NB85 | - | 28.00 | PI | TERESINA | 6-2022 | F | 24702 | 86 | 1847.9 | 96.4 | 97.7 |
| CHIKV\|PI\|NB86\|2022-06 | NB86 | - | 23.00 | PI | TERESINA | 6-2022 | M | 17699 | 92.9 | 1273.1 | 96.4 | 97.9 |
| CHIKV\|PI\|NB87\|2022-06 | NB87 | - | 28.00 | PI | DEMERVAL LOBÃO | 6-2022 | F | 17376 | 86.6 | 1222 | 96.4 | 97.8 |
| CHIKV\|PI\|NB88\|2022-06 | NB88 | - | 28.00 | PI | TERESINA | 6-2022 | F | 17382 | 86.3 | 1363.8 | 96.4 | 97.7 |
| CHIKV\|PI\|NB89\|2022-06 | NB89 | - | 30.00 | PI | TERESINA | 6-2022 | F | 34966 | 85.7 | 2286.7 | 96.5 | 97.8 |
| CHIKV\|PI\|NB90\|2022-06 | NB90 | - | 28.00 | PI | TERESINA | 6-2022 | M | 46180 | 85 | 2683.8 | 96.3 | 97.5 |
| CHIKV\|PI\|NB91\|2022-06 | NB91 | - | 26.00 | PI | TERESINA | 6-2022 | F | 26429 | 91.9 | 2069.9 | 96.6 | 98.1 |
| CHIKV\|NBC01\|PR199_2096-21\|2021-04 | BC1 | 2096/21 | 32.00 | PR | APUCARANA | 4-2021 | M | 151883 | 61.1 | 11994.1 | - | - |
| CHIKV\|NBC02\|PR200_2221-21\|2021-04 | BC2 | 2221/21 | 28.00 | PR | APUCARANA | 4-2021 | M | 160235 | 69.1 | 39235.6 | - | - |
| CHIKV\|NBC03\|PR201_2250-21\|2021-04 | BC3 | 2250/21 | 25.00 | PR | APUCARANA | 4-2021 | F | 167809 | 63 | 16195.3 | - | - |
| CHIKV\|NBC04\|PR203_2254-21\|2021-04 | BC4 | 2254/21 | 28.00 | PR | APUCARANA | 4-2021 | M | 237806 | 66.5 | 18348.6 | - | - |
| CHIKV\|NBC12\|1510-21\|PR\|2021-03 | BC12 | 1510/21 | 28.00 | PR | APUCARANA | 3-2021 | F | 9533 | 85.1 | 626.2 | 96.8 | 98.1 |
| CHIKV\|NBC13\|1885-21\|PR\|2021-04 | BC13 | 1885/21 | 25.00 | PR | APUCARANA | 4-2021 | F | 11817 | 85.6 | 846.7 | 96.9 | 98.1 |
| CHIKV\|NBC14\|1886-21\|PR\|2021-04 | BC14 | 1886/21 | 27.00 | PR | APUCARANA | 4-2021 | F | 16728 | 84.1 | 1088 | 96.9 | 97.9 |
| CHIKV\|NBC15_2094-21\|PR\|2021-04 | BC15 | 2094/21 | 30.00 | PR | APUCARANA | 4-2021 | F | 8409 | 74.2 | 514.4 | 96.7 | 97.7 |
| CHIKV\|NBC16_2049-21\|PR\|2021-04 | BC16 | 2049/21 | 24.00 | PR | APUCARANA | 4-2021 | F | 11043 | 82.5 | 755.7 | 96.9 | 98.1 |
| CHIKV\|NBC17\|1204-21\|PR\|2021-03 | BC17 | 1204/21 | 25.00 | PR | APUCARANA | 3-2021 | M | 10362 | 84.8 | 723.3 | 96.9 | 98.2 |
| CHIKV\|NBC18\|1406-21\|PR\|2021-03 | BC18 | 1406/21 | 33.00 | PR | APUCARANA | 3-2021 | F | 1854 | 55.5 | 180.8 | 96.6 | 97.5 |
| CHIKV\|NBC19\|1290-21\|PR\|2021-03 | BC19 | 1290/21 | 29.00 | PR | APUCARANA | 3-2021 | F | 10815 | 78.8 | 736.8 | 96.8 | 98 |
| CHIKV\|NBC20\|1984-21\|PR\|2021-04 | BC20 | 1984/21 | 30.00 | PR | APUCARANA | 4-2021 | F | 11587 | 81.2 | 671.2 | 96.9 | 98 |
| CHIKV\|NBC21\|1439-21\|PR\|2021-03 | BC21 | 1439/21 | 29.00 | PR | APUCARANA | 3-2021 | F | 21438 | 82.1 | 1377.4 | 96.9 | 98 |
| CHIKV\|RN\|ZDC-933\|2022-04 | BC22 | ZDC 933 | 25.00 | RN | NOVA CRUZ | 4-2022 | M | 22831 | 94 | 1903.12 | 96.6 | 98.2 |

| | | | | | | | | | | | | |
|---|---|---|---|---|---|---|---|---|---|---|---|---|
| CHIKV\|RN\|ZDC-719\|2022-03 | BC49 | ZDC 719 | 29.00 | RN | PARELHAS | 3-2022 | M | 13726 | 93.3 | 808.5 | 96.7 | 98.2 |
| CHIKV\|RN\|ZDC-720\|2022-03 | BC50 | ZDC 720 | 30.00 | RN | PARELHAS | 3-2022 | M | 14267 | 85.7 | 886.8 | 96.5 | 98 |
| CHIKV\|RN\|ZDC-723\|2022-03 | BC51 | ZDC 723 | 23.00 | RN | PARELHAS | 3-2022 | F | 16660 | 94.4 | 1403.1 | 96.5 | 98 |
| CHIKV\|RN\|ZDC-790\|2022-04 | BC52 | ZDC 790 | 22.00 | RN | POCO BRANCO | 4-2022 | F | 75876 | 93.6 | 5957.9 | 96.7 | 98.3 |
| CHIKV\|RN\|ZDC-796\|2022-03 | BC53 | ZDC 796 | 31.00 | RN | ACARI | 3-2022 | F | 34090 | 94.4 | 2313.8 | 96.6 | 98.2 |
| CHIKV\|RN\|ZDC-798\|2022-03 | BC54 | ZDC 798 | 25.00 | RN | ACARI | 3-2022 | M | 33333 | 94.6 | 2107.7 | 96.6 | 98.2 |
| CHIKV\|RN\|ZDC-816\|2022-04 | BC55 | ZDC 816 | 23.00 | RN | TIBAU DO SUL | 4-2022 | M | 37289 | 94.4 | 2756.6 | 96.4 | 98 |
| CHIKV\|RN\|ZDC-820\|2022-04 | BC56 | ZDC 820 | 23.00 | RN | JUNDIA | 4-2022 | M | 46488 | 94.1 | 2939.4 | 96.6 | 98.2 |
| CHIKV\|RN\|ZDC-822\|2022-04 | BC57 | ZDC 822 | 28.00 | RN | JUNDIA | 4-2022 | F | 18565 | 92.2 | 1045 | 96.7 | 98.1 |
| CHIKV\|RN\|ZDC-825\|2022-04 | BC58 | ZDC 825 | 25.00 | RN | SANTA CRUZ | 4-2022 | F | 24725 | 94.5 | 1617.2 | 96.6 | 98.3 |
| CHIKV\|RN\|ZDC-826\|2022-04 | BC59 | ZDC 826 | 25.00 | RN | SANTA CRUZ | 4-2022 | M | 21229 | 93.6 | 1376.7 | 96.6 | 98.2 |
| CHIKV\|RN\|ZDC-829\|2022-04 | BC60 | ZDC 829 | 23.00 | RN | MACAIBA | 4-2022 | F | 24580 | 94.6 | 1623.6 | 96.5 | 98.2 |
| CHIKV\|RN\|ZDC-845\|2022-04 | BC61 | ZDC 845 | 19.00 | RN | NOVA CRUZ | 4-2022 | M | 23434 | 94.1 | 1731.5 | 96.6 | 98.2 |
| CHIKV\|RN\|ZDC-932\|2022-04 | BC62 | ZDC 932 | 19.00 | RN | NOVA CRUZ | 4-2022 | F | 36594 | 94.7 | 3179.1 | 96.5 | 98.1 |
| CHIKV\|RN\|ZDC-938\|2022-04 | BC63 | ZDC 938 | 23.00 | RN | NOVA CRUZ | 4-2022 | F | 30512 | 94.2 | 2046.9 | 96.6 | 98.1 |
| CHIKV\|RN\|ZDC-939\|2022-04 | BC64 | ZDC 939 | 24.00 | RN | NOVA CRUZ | 4-2022 | F | 28374 | 94.6 | 1703.3 | 96.3 | 98 |
| CHIKV\|RN\|ZDC-968\|2022-04 | BC65 | ZDC 968 | 23.00 | RN | JANDAIRA | 4-2022 | M | 32854 | 94.6 | 2161.3 | 96.6 | 98.1 |
| CHIKV\|RN\|ZDC-1035\|2022-04 | BC66 | ZDC 1035 | 29.00 | RN | PARNAMIRIM | 4-2022 | F | 19157 | 91.3 | 1155.5 | 96.4 | 98.1 |
| CHIKV\|RN\|ZDC-1038\|2022-05 | BC67 | ZDC 1038 | 24.00 | RN | NATAL | 5-2022 | F | 20378 | 93.2 | 1325.4 | 96.7 | 98.2 |
| CHIKV\|RN\|ZDC-1119\|2022-04 | BC68 | ZDC 1119 | 22.00 | RN | SANTA CRUZ | 4-2022 | M | 28747 | 94.1 | 2151.1 | 96.7 | 98.3 |
| CHIKV\|RN\|ZDC-1123\|2022-05 | BC69 | ZDC 1123 | 23.00 | RN | TIBAU DO SUL | 5-2022 | F | 36601 | 94.4 | 2324 | 96.6 | 98.2 |
| CHIKV\|RN\|ZDC-1125\|2022-05 | BC70 | ZDC 1125 | 19.00 | RN | PARNAMIRIM | 5-2022 | M | 48868 | 95 | 3424.7 | 96.5 | 98.1 |
| CHIKV\|RN\|ZDC-1170\|2022-05 | BC71 | ZDC 1170 | 28.00 | RN | MACAU | 5-2022 | F | 27905 | 94.2 | 1667.4 | 96.7 | 98.2 |
| CHIKV\|RN\|ZDC-1237\|2022-05 | BC72 | ZDC 1237 | 25.00 | RN | SITIO NOVO | 5-2022 | F | 14196 | 89.9 | 871 | 96.6 | 98.1 |
| CHIKV\|RN\|ZDC-1293\|2022-05 | BC73 | ZDC 1293 | 21.00 | RN | JANDAIRA | 5-2022 | F | 31397 | 94.7 | 1889.4 | 96.6 | 98.1 |
| CHIKV\|RN\|ZDC-1294\|2022-05 | BC74 | ZDC 1294 | 17.00 | RN | JANDAIRA | 5-2022 | M | 33118 | 94.5 | 2512.2 | 96.6 | 98.2 |
| CHIKV\|RN\|ZDC-1305\|2022-05 | BC75 | ZDC 1305 | 23.00 | RN | SANTO ANTONIO | 5-2022 | M | 26277 | 93 | 1628.9 | 96.7 | 98.1 |
| CHIKV\|RN\|ZDC-1362\|2022-05 | BC76 | ZDC 1362 | 27.00 | RN | MONTE ALEGRE | 5-2022 | M | 30405 | 94.1 | 2037.6 | 96.6 | 98.2 |
| CHIKV\|RN\|ZDC-1367\|2022-05 | BC77 | ZDC 1367 | 20.00 | RN | MONTE ALEGRE | 5-2022 | F | 38156 | 92.9 | 2430.5 | 96.6 | 98.1 |
| CHIKV\|RN\|ZDC-1368\|2022-05 | BC78 | ZDC 1368 | 21.00 | RN | MONTE ALEGRE | 5-2022 | F | 34587 | 94 | 2158.8 | 96.7 | 98.3 |
| CHIKV\|RN\|ZDC-1369\|2022-05 | BC79 | ZDC 1369 | 16.00 | RN | MONTE ALEGRE | 5-2022 | F | 14418 | 94.3 | 1045.7 | 96.7 | 98.3 |
| CHIKV\|RN\|ZDC-1418\|2022-05 | BC81 | ZDC 1418 | 17.00 | RN | IELMO MARINHO | 5-2022 | F | 21005 | 94 | 1303.7 | 96.6 | 98.2 |
| CHIKV\|RN\|ZDC-1485\|2022-05 | BC82 | ZDC 1485 | 23.00 | RN | JOAO CAMARA | 5-2022 | F | 31660 | 94.2 | 2081.3 | 96.6 | 98.1 |

| | | | | | | | | | | | | |
|---|---|---|---|---|---|---|---|---|---|---|---|---|
| CHIKV\|RN\|ZDC-1492\|2022-05 | BC83 | ZDC 1492 | 21.00 | RN | JOAO CAMARA | 5-2022 | M | 33367 | 94.1 | 2062 | 96.7 | 98.2 |
| CHIKV\|RN\|ZDC-1495\|2022-05 | BC84 | ZDC 1495 | 19.00 | RN | JOAO CAMARA | 5-2022 | M | 38372 | 94.1 | 2323.5 | 96.7 | 98.2 |
| CHIKV\|RN\|ZDC-1457\|2022-05 | BC85 | ZDC 1457 | 24.00 | RN | PARNAMIRIM | 5-2022 | F | 31129 | 94.5 | 1902.2 | 96.6 | 98.2 |
| CHIKV\|RN\|ZDC-1300\|2022-05 | BC86 | ZDC 1300 | 25.00 | RN | JANDAIRA | 5-2022 | F | 15458 | 93.6 | 937.4 | 96.8 | 98.2 |
| CHIKV\|RN\|ZDC-1505\|2022-05 | BC87 | ZDC 1505 | 18.00 | RN | NATAL | 5-2022 | M | 24821 | 92.1 | 1358.2 | 96.5 | 97.9 |
| CHIKV\|RN\|ZDC-1529\|2022-05 | BC88 | ZDC 1529 | 20.00 | RN | JANDAIRA | 5-2022 | M | 24643 | 93.7 | 1514.1 | 96.7 | 98.1 |
| CHIKV\|RN\|ZDC-1607\|2022-05 | BC89 | ZDC 1607 | 21.00 | RN | SANTA MARIA | 5-2022 | M | 24053 | 94.3 | 1466.8 | 96.7 | 98.3 |
| CHIKV\|RN\|ZDC-2129\|2022-06 | BC90 | ZDC 2129 | 26.00 | RN | NATAL | 6-2022 | F | 18897 | 92.6 | 1145.4 | 96.8 | 98.2 |
| CHIKV\|RN\|ZDC-2131\|2022-06 | BC91 | ZDC 2131 | 21.00 | RN | NATAL | 6-2022 | F | 21180 | 94.8 | 1407 | 96.5 | 98.1 |
| CHIKV\|RN\|1945\|2022-05 | NB93 | 1945 (Natal) | - | RN | Acari | 5-2022 | M | 40492 | 90.3 | 2465.1 | 96.5 | 98 |
| CHIKV\|RN\|1826\|2022-05 | NB94 | 1826 (Natal) | - | RN | Nova Cruz | 5-2022 | F | 19252 | 62.9 | 1494.8 | 95.9 | 96.9 |
| CHIKV\|NB45\|SE-01\|2021-09 | NB45 | SE-01 | 21.33 | SE | INDIAROBA | 9-2021 | F | 1880 | 93 | 128.5 | 96.9 | 98.4 |
| CHIKV\|NB46\|SE-02\|2021-09 | NB46 | SE-02 | 18.00 | SE | PORTO DA FOLHA | 9-2021 | M | 110248 | 94.3 | 8344.5 | 96.7 | 98.2 |
| CHIKV\|NB47\|SE-03\|2021-09 | NB47 | SE-03 | 18.51 | SE | PORTO DA FOLHA | 9-2021 | M | 106318 | 94.2 | 7110.1 | 96.7 | 98.2 |
| CHIKV\|NB48\|SE-04\|2021-09 | NB48 | SE-04 | 18.14 | SE | NEOPOLIS | 9-2021 | F | 94148 | 93.6 | 7163.6 | 96.6 | 98 |
| CHIKV\|NB49\|SE-05\|2021-09 | NB49 | SE-05 | 22.00 | SE | INDIAROBA | 9-2021 | F | 118972 | 93.5 | 6326.2 | 96.7 | 98.1 |
| CHIKV\|NB50\|SE-06\|2021-09 | NB50 | SE-06 | 22.34 | SE | PORTO DA FOLHA | 9-2021 | F | 135111 | 94.5 | 8731.7 | 96.7 | 98.2 |
| CHIKV\|NB51\|SE-07\|2021-09 | NB51 | SE-07 | 19.46 | SE | LAGARTO | 9-2021 | M | 100335 | 95 | 7164.9 | 96.4 | 98.1 |
| CHIKV\|NB52\|SE-08\|2021-09 | NB52 | SE-08 | 18.61 | SE | LAGARTO | 9-2021 | F | 52198 | 92.6 | 3633.4 | 96.8 | 98.3 |
| CHIKV\|NB53\|SE-09\|2021-09 | NB53 | SE-09 | 22.45 | SE | LAGARTO | 9-2021 | M | 73612 | 94 | 5180.4 | 96.7 | 98.2 |
| CHIKV\|NB54\|SE-10\|2021-09 | NB54 | SE-10 | 14.00 | SE | ARACAJU | 9-2021 | F | 106552 | 94.8 | 9112.9 | 96.7 | 98.1 |
| CHIKV\|NB55\|SE-11\|2021-09 | NB55 | SE-11 | 23.08 | SE | SAO CRISTOVAO | 9-2021 | M | 101483 | 93.2 | 5573 | 96.8 | 98.2 |
| CHIKV\|NB56\|SE-12\|2021-09 | NB56 | SE-12 | 21.00 | SE | PORTO DA FOLHA | 9-2021 | F | 96469 | 94.6 | 6281.7 | 96.7 | 98.2 |
| CHIKV\|NB57\|SE-13\|2021-09 | NB57 | SE-13 | 21.07 | SE | PORTO DA FOLHA | 9-2021 | F | 51279 | 94.1 | 3802.4 | 96.7 | 98.2 |
| CHIKV\|NB58\|SE-14\|2021-09 | NB58 | SE-14 | 19.00 | SE | PORTO DA FOLHA | 9-2021 | F | 98971 | 95.5 | 6908.8 | 96.3 | 98.3 |
| CHIKV\|NB59\|SE-15\|2021-09 | NB59 | SE-15 | 19.00 | SE | INDIAROBA | 9-2021 | F | 58527 | 93.9 | 4795.6 | 96.7 | 98.2 |
| CHIKV\|NB60\|SE-16\|2021-09 | NB60 | SE-16 | 20.00 | SE | NEOPOLIS | 9-2021 | F | 49195 | 94.2 | 3461 | 96.7 | 98.3 |
| CHIKV\|NB61\|SE-17\|2021-09 | NB61 | SE-17 | 19.49 | SE | NEOPOLIS | 9-2021 | F | 19353 | 93.3 | 1599.9 | 96.7 | 98.2 |
| CHIKV\|NB62\|SE-18\|2021-09 | NB62 | SE-18 | 21.00 | SE | PORTO DA FOLHA | 9-2021 | M | 32695 | 94.6 | 2492.5 | 96.6 | 98.1 |
| CHIKV\|NB63\|SE-19\|2021-09 | NB63 | SE-19 | 18.77 | SE | PORTO DA FOLHA | 9-2021 | F | 58053 | 94.1 | 4533.8 | 96.7 | 98.2 |
| CHIKV\|NB64\|SE-20\|2021-09 | NB64 | SE-20 | 22.26 | SE | PORTO DA FOLHA | 9-2021 | M | 63387 | 95 | 4991.5 | 96.5 | 98.2 |
| CHIKV\|NB65\|SE-21\|2021-09 | NB65 | SE-21 | 21.75 | SE | NEOPOLIS | 9-2021 | F | 117427 | 94.8 | 7486.4 | 96.5 | 98 |
| CHIKV\|NB66\|SE-22\|2021-10 | NB66 | SE-22 | 24.63 | SE | UMBAUBA | 10-2021 | F | 34284 | 94.5 | 2208.5 | 96.6 | 98.2 |

| | | | | | | | | | | | | |
|---|---|---|---|---|---|---|---|---|---|---|---|---|
| CHIKV\|NB67\|SE-23\|2021-09 | NB67 | SE-23 | 20.21 | SE | LAGARTO | 9-2021 | F | 52885 | 92.3 | 3639.7 | 96.8 | 98.2 |
| CHIKV\|NB68\|SE-24\|2021-09 | NB68 | SE-24 | 22.17 | SE | LAGARTO | 9-2021 | M | 25638 | 94 | 1685.2 | 96.7 | 98.2 |
| CHIKV\|NB69\|SE-25\|2021-10 | NB69 | SE-25 | 17.96 | SE | NEOPOLIS | 10-2021 | F | 65546 | 95.5 | 5133.3 | 96.3 | 98.2 |
| CHIKV\|NB70\|SE-26\|2021-10 | NB70 | SE-26 | 21.58 | SE | PORTO DA FOLHA | 10-2021 | M | 21223 | 92.9 | 1228.6 | 96.8 | 98.1 |
| CHIKV\|NB71\|SE-27\|2021-10 | NB71 | SE-27 | 20.18 | SE | PORTO DA FOLHA | 10-2021 | M | 2253 | 92.1 | 132.7 | 96.8 | 98.3 |
| CHIKV\|NB72\|SE-28\|2021-10 | NB72 | SE-28 | 21.58 | SE | NEOPOLIS | 10-2021 | F | 52344 | 93.5 | 3745.6 | 96.7 | 98.2 |
| CHIKV\|NB73\|SE-29\|2021-10 | NB73 | SE-29 | 20.17 | SE | PORTO DA FOLHA | 10-2021 | M | 61864 | 94.2 | 4416.5 | 96.7 | 98.2 |
| CHIKV\|NB74\|SE-30\|2021-09 | NB74 | SE-30 | 21.23 | SE | INDIAROBA | 9-2021 | M | 62198 | 94.8 | 4092.9 | 96.5 | 97.9 |
| CHIKV\|NB75\|SE-31\|2021-10 | NB75 | SE-31 | 18.32 | SE | PORTO DA FOLHA | 10-2021 | F | 100856 | 93.6 | 6944.3 | 96.6 | 98 |
| CHIKV\|NB76\|SE-32\|2021-10 | NB76 | SE-32 | 22.00 | SE | PORTO DA FOLHA | 10-2021 | F | 44672 | 92.3 | 3317.7 | 96.7 | 98.2 |
| CHIKV\|NB77\|SE-33\|2021-10 | NB77 | SE-33 | 16.00 | SE | MONTE ALEGRE DE SERGIPE | 10-2021 | M | 77541 | 94.2 | 6055.2 | 96.7 | 98.2 |
| CHIKV\|NB78\|SE-34\|2021-10 | NB78 | SE-34 | 24.32 | SE | LAGARTO | 10-2021 | M | 90753 | 93.3 | 6368.9 | 96.7 | 98.2 |
| CHIKV\|NB79\|SE-35\|2021-10 | NB79 | SE-35 | 19.53 | SE | PORTO DA FOLHA | 10-2021 | F | 45355 | 93.2 | 3198.8 | 96.7 | 98.2 |
| CHIKV\|NB80\|SE-36\|2021-10 | NB80 | SE-36 | 19.92 | SE | PORTO DA FOLHA | 10-2021 | M | 48604 | 93.2 | 3189.4 | 96.7 | 98.3 |
| CHIKV\|NB81\|SE-37\|2021-10 | NB81 | SE-37 | 23.47 | SE | ARACAJU | 10-2021 | M | 2661 | 94.1 | 138 | 96.8 | 98.3 |
| CHIKV\|NB82\|SE-38\|2021-10 | NB82 | SE-38 | 14.98 | SE | MONTE ALEGRE DE SERGIPE | 10-2021 | F | 2433 | 94.3 | 119.3 | 96.8 | 98.4 |
| CHIKV\|NB83\|SE-39\|2021-10 | NB83 | SE-39 | 19.06 | SE | PORTO DA FOLHA | 10-2021 | M | 2726 | 94.1 | 126.2 | 96.8 | 98.3 |
| CHIKV\|NB84\|SE-40\|2021-10 | NB84 | SE-40 | 22.96 | SE | PORTO DA FOLHA | 10-2021 | M | 2893 | 92.4 | 171.6 | 96.8 | 98.3 |
| CHIKV\|NB85\|SE-41\|2021-10 | NB85 | SE-41 | 20.55 | SE | PORTO DA FOLHA | 10-2021 | F | 2214 | 92.7 | 136.9 | 96.8 | 98.3 |
| CHIKV\|NB86\|SE-42\|2021-10 | NB86 | SE-42 | 19.40 | SE | PORTO DA FOLHA | 10-2021 | M | 2937 | 92.9 | 158.7 | 96.8 | 98.3 |
| CHIKV\|NB87\|SE-43\|2021-10 | NB87 | SE-43 | 21.97 | SE | PORTO DA FOLHA | 10-2021 | F | 1971 | 92.4 | 107.1 | 96.9 | 98.3 |
| CHIKV\|NB88\|SE-44\|2021-10 | NB88 | SE-44 | 32.36 | SE | SIMAO DIAS | 10-2021 | F | 2609 | 93.9 | 148.4 | 96.8 | 98.3 |
| CHIKV\|MG\|25-22-RED\|2021-12 | UDP0318 | 25.22_RED | 22.70 | MG | GOVERNADOR VALADARES | 12-2021 | F | 941173 | 96 | 8767.19 | - | - |
| CHIKV\|MG\|270-22-RED\|2022-02 | UDP0319 | 270.22_RED | 25.80 | MG | MONTES CLAROS | 2-2022 | F | 835235 | 95 | 7554.2 | - | - |
| CHIKV\|MG\|571-22-RED\|2022-03 | UDP0320 | 571.22_RED | 22.50 | MG | FRANCISCO SA | 3-2022 | M | 870644 | 96 | 8020.93 | - | - |
| CHIKV\|MG\|612-22-RED\|2022-02 | UDP0321 | 612.22_RED | 26.20 | MG | BELO HORIZONTE | 2-2022 | F | 906493 | 96 | 8369.2 | - | - |
| CHIKV\|MG\|726-22-RED\|2022-03 | UDP0322 | 726.22_RED | 23.40 | MG | SALINAS | 3-2022 | M | 364403 | 96 | 3326.73 | - | - |
| CHIKV\|MG\|736-22-RED\|2022-03 | UDP0323 | 736.22_RED | 34.00 | MG | MONTE AZUL | 3-2022 | F | 188645 | 42 | 1612.93 | - | - |
| CHIKV\|MG\|1754-22-RED\|2022-03 | UDP0324 | 1754.22_RED | 24.40 | MG | ARACUAI | 3-2022 | F | 892530 | 96 | 8247.74 | - | - |
| CHIKV\|MG\|768-22-C-RED\|2022-03 | UDP0325 | 768.22_C_RED | 28.50 | MG | MONTES CLAROS | 3-2022 | M | 1011157 | 96 | 9348.75 | - | - |
| CHIKV\|BA\|BC01\|2022-03 | BC01 | 292346970 | 21.30 | BA | ITAJUIPE | 3-2022 | F | 14403.0 | 94.9 | 1024.3 | 96.4 | 97.9 |
| CHIKV\|BA\|BC02\|2022-03 | BC02 | 292342712 | 30.40 | BA | ITAJUIPE | 3-2022 | F | 21384.0 | 87.5 | 1171.4 | 96.4 | 97.7 |
| CHIKV\|BA\|BC03\|2022-03 | BC03 | 292342523 | 21.10 | BA | ITAJUIPE | 3-2022 | F | 23675.0 | 94.4 | 1539.3 | 96.7 | 98.2 |

| | | | | | | | | | | | | |
|---|---|---|---|---|---|---|---|---|---|---|---|---|
| CHIKV\|BA\|BC04\|2022-03 | BC04 | 292347014 | 24.60 | BA | ITAJUIPE | 3-2022 | M | 29038.0 | 94.0 | 1559.7 | 96.7 | 98.2 |
| CHIKV\|BA\|BC05\|2022-03 | BC05 | 292342731 | 23.60 | BA | ITAJUIPE | 3-2022 | F | 24475.0 | 94.4 | 1462.5 | 96.6 | 98.0 |
| CHIKV\|BA\|BC06\|2022-02 | BC06 | 292318861 | 25.60 | BA | ITAJUIPE | 2-2022 | M | 42181.0 | 94.3 | 2232.2 | 96.5 | 98.0 |
| CHIKV\|BA\|BC07\|2022-03 | BC07 | 292340189 | 21.90 | BA | JUAZEIRO | 3-2022 | M | 24179.0 | 94.1 | 1619.3 | 96.8 | 98.2 |
| CHIKV\|BA\|BC08\|2022-03 | BC08 | 292309317 | 28.60 | BA | ITAPETINGA | 3-2022 | F | 25283.0 | 87.8 | 1285.2 | 96.8 | 98.2 |
| CHIKV\|BA\|BC09\|2022-03 | BC09 | 292329398 | 22.00 | BA | ITUIUTABA | 3-2022 | M | 1735.0 | 93.1 | 112.5 | 96.8 | 98.1 |
| CHIKV\|BA\|BC10\|2022-03 | BC10 | 292345418 | 24.50 | BA | JACOBINA | 3-2022 | F | 27959.0 | 94.0 | 1740.2 | 96.7 | 98.2 |
| CHIKV\|BA\|BC11\|2022-03 | BC11 | 292341930 | 21.60 | BA | MADRE DE DEUS | 3-2022 | F | 27264.0 | 94.1 | 1919.2 | 96.7 | 98.2 |
| CHIKV\|BA\|BC12\|2022-03 | BC12 | 292339837 | 23.50 | BA | UAUA | 3-2022 | M | 31843.0 | 94.7 | 1863.1 | 96.7 | 98.2 |
| CHIKV\|BA\|BC13\|2022-03 | BC13 | 292354609 | 17.90 | BA | ALMADINA | 3-2022 | F | 26158.0 | 94.5 | 2001.9 | 96.6 | 98.1 |
| CHIKV\|BA\|BC14\|2022-03 | BC14 | 292342338 | 20.30 | BA | COARACI | 3-2022 | F | 28528.0 | 94.3 | 1834.3 | 96.6 | 98.2 |
| CHIKV\|BA\|BC15\|2022-03 | BC15 | 292324218 | 16.40 | BA | COARACI | 3-2022 | M | 17553.0 | 94.4 | 1366.6 | 96.7 | 98.2 |
| CHIKV\|BA\|BC16\|2022-03 | BC16 | 292324308 | 27.30 | BA | COARACI | 3-2022 | F | 27338.0 | 94.7 | 1537.5 | 96.4 | 98.1 |
| CHIKV\|BA\|BC17\|2022-03 | BC17 | 292335413 | 28.60 | BA | FEIRA DE SANTANA | 3-2022 | F | 6640.0 | 89.0 | 353.8 | 96.6 | 97.8 |
| CHIKV\|BA\|BC18\|2022-03 | BC18 | 292351417 | 26.90 | BA | ITAJUIPE | 3-2022 | M | 16125.0 | 94.0 | 1251.3 | 96.6 | 98.2 |
| CHIKV\|BA\|BC19\|2022-03 | BC19 | 292342764 | 27.90 | BA | ITAJUIPE | 3-2022 | F | 22548.0 | 94.1 | 1109.0 | 96.7 | 98.2 |
| CHIKV\|BA\|BC20\|2022-03 | BC20 | 292342653 | 27.20 | BA | ITAJUIPE | 3-2022 | M | 29063.0 | 92.9 | 1603.0 | 96.7 | 98.1 |
| CHIKV\|BA\|BC21\|2022-03 | BC21 | 292326836 | 18.90 | BA | ITAJUIPE | 3-2022 | M | 33825.0 | 94.5 | 2267.3 | 96.6 | 98.1 |
| CHIKV\|BA\|BC22\|2022-03 | BC22 | 292326999 | 22.70 | BA | ITAJUIPE | 3-2022 | F | 33317.0 | 94.0 | 2031.0 | 96.6 | 98.1 |
| CHIKV\|BA\|BC23\|2022-03 | BC23 | 292326262 | 26.90 | BA | ITAMARAJU | 3-2022 | M | 30156.0 | 92.6 | 1658.3 | 96.7 | 98.1 |
| CHIKV\|BA\|BC24\|2022-03 | BC24 | 292341330 | 26.00 | BA | ITAPITANGA | 3-2022 | F | 33698.0 | 94.1 | 1862.7 | 96.4 | 98.1 |
| CHIKV\|BA\|BC25\|2022-03 | BC25 | 292333190 | 21.80 | BA | JACOBINA | 3-2022 | F | 8149.0 | 94.4 | 604.9 | 96.6 | 98.0 |
| CHIKV\|BA\|BC26\|2022-03 | BC26 | 292329319 | 27.80 | BA | JUAZEIRO | 3-2022 | M | 25549.0 | 94.5 | 1315.3 | 96.7 | 98.2 |
| CHIKV\|BA\|BC27\|2022-05 | BC27 | 292444774 | 23.60 | BA | PARIPIRANGA | 5-2022 | F | 20410.0 | 94.5 | 1243.8 | 96.7 | 98.1 |
| CHIKV\|BA\|BC28\|2022-05 | BC28 | 292444767 | 20.10 | BA | PARIPIRANGA | 5-2022 | M | 20000.0 | 94.1 | 1425.5 | 96.7 | 98.2 |
| CHIKV\|BA\|BC29\|2022-05 | BC29 | 292442988 | 20.80 | BA | JEQUIE | 5-2022 | F | 25754.0 | 94.3 | 1585.8 | 96.6 | 98.1 |
| CHIKV\|BA\|BC30\|2022-05 | BC30 | 292452959 | 18.70 | BA | ITABUNA | 5-2022 | F | 20391.0 | 94.1 | 1467.5 | 96.7 | 98.2 |
| CHIKV\|BA\|BC31\|2022-06 | BC31 | 292456977 | 16.60 | BA | POJUCA | 6-2022 | F | 36710.0 | 93.6 | 2164.9 | 96.7 | 98.1 |
| CHIKV\|BA\|BC32\|2022-06 | BC32 | 292468683 | 19.80 | BA | CAMACARI | 6-2022 | M | 25695.0 | 94.5 | 1683.7 | 96.7 | 98.3 |
| CHIKV\|BA\|BC33\|2022-05 | BC33 | 292419147 | 20.90 | BA | ITABUNA | 5-2022 | F | 20607.0 | 94.2 | 1346.2 | 96.6 | 98.1 |
| CHIKV\|BA\|BC34\|2022-05 | BC34 | 292419397 | 17.40 | BA | ITABUNA | 5-2022 | M | 14290.0 | 94.7 | 1098.7 | 96.6 | 98.1 |
| CHIKV\|BA\|BC35\|2022-05 | BC35 | 292419463 | 21.30 | BA | ITABUNA | 5-2022 | F | 28251.0 | 92.3 | 1661.1 | 96.7 | 98.0 |
| CHIKV\|BA\|BC36\|2022-04 | BC36 | 292399973 | 22.30 | BA | ITABUNA | 4-2022 | F | 31974.0 | 94.4 | 1927.8 | 96.4 | 97.9 |

| | | | | | | | | | | | | |
|---|---|---|---|---|---|---|---|---|---|---|---|---|
| CHIKV\|BA\|BC37\|2022-04 | BC37 | 292397013 | 21.00 | BA | ITAJUIPE | 4-2022 | F | 28691.0 | 94.4 | 1825.3 | 96.6 | 98.1 |
| CHIKV\|BA\|BC38\|2022-04 | BC38 | 292373156 | 21.50 | BA | ITAJUIPE | 4-2022 | M | 30704.0 | 93.9 | 2297.8 | 96.7 | 98.2 |
| CHIKV\|BA\|BC39\|2022-04 | BC39 | 292385170 | 29.90 | BA | JUAZEIRO | 4-2022 | M | 14959.0 | 88.3 | 808.1 | 96.7 | 98.2 |
| CHIKV\|BA\|BC40\|2022-04 | BC40 | 292395603 | 20.30 | BA | EUNAPOLIS | 4-2022 | F | 24366.0 | 94.0 | 1604.8 | 96.7 | 98.2 |
| CHIKV\|BA\|BC41\|2022-04 | BC41 | 292397361 | 17.80 | BA | ILHEUS | 4-2022 | M | 16506.0 | 94.1 | 1287.3 | 96.6 | 98.2 |
| CHIKV\|BA\|BC42\|2022-04 | BC42 | 292395830 | 18.70 | BA | ITABUNA | 4-2022 | F | 18083.0 | 94.2 | 1249.9 | 96.6 | 98.2 |
| CHIKV\|BA\|BC43\|2022-05 | BC43 | 292408228 | 17.50 | BA | ITABUNA | 5-2022 | M | 20984.0 | 94.1 | 1646.4 | 96.6 | 98.2 |
| CHIKV\|BA\|BC44\|2022-04 | BC44 | 292398529 | 20.70 | BA | MADRE DE DEUS | 4-2022 | M | 22470.0 | 94.6 | 1469.8 | 96.7 | 98.2 |
| CHIKV\|BA\|BC45\|2022-04 | BC45 | 292378738 | 23.20 | BA | MORRO DO CHAPEU | 4-2022 | M | 28955.0 | 93.5 | 1730.6 | 96.7 | 98.1 |
| CHIKV\|BA\|BC46\|2022-04 | BC46 | 292397438 | 21.20 | BA | SERRA | 4-2022 | M | 23831.0 | 94.4 | 1500.1 | 96.6 | 98.1 |
| CHIKV\|BA\|BC47\|2022-04 | BC47 | 292370309 | 19.80 | BA | CAMPO FORMOSO | 4-2022 | M | 23126.0 | 94.3 | 1597.5 | 96.7 | 98.2 |
| CHIKV\|BA\|BC48\|2022-03 | BC48 | 292316064 | 35.90 | BA | ITAMARAJU | 3-2022 | F | 23841.0 | 78.7 | 1476.4 | 96.4 | 98.0 |
| CHIKV\|BA\|BC94\|2022-03 | BC94 | 292350702 | 23.60 | BA | PORTO SEGURO | 3-2022 | F | 25149.0 | 91.6 | 1256.5 | 96.7 | 98.2 |
| CHIKV\|BA\|FS0030\|2015-07 | NB35 | FS0030 | 26.00 | BA | Feira de Santana | 7-2015 | F | 12212.0 | 73.8 | 780.6 | 96.4 | 97.8 |
| CHIKV\|BA\|FS0032\|2015-07 | NB36 | FS0032 | 27.00 | BA | Feira de Santana | 7-2015 | F | 25812.0 | 77.6 | 1401.0 | 96.8 | 98.1 |
| CHIKV\|BA\|FS0035\|2015-07 | NB37 | FS0035 | 23.00 | BA | Feira de Santana | 7-2015 | M | 23109.0 | 90.6 | 1099.6 | 96.8 | 98.2 |
| CHIKV\|BA\|FS0047\|2015-07 | NB38 | FS0047 | 28.00 | BA | Feira de Santana | 7-2015 | F | 14481.0 | 73.8 | 889.1 | 96.8 | 98.0 |
| CHIKV\|BA\|FS0048\|2015-07 | NB39 | FS0048 | 30.00 | BA | Feira de Santana | 7-2015 | M | 8604.0 | 59.1 | 621.6 | 96.8 | 97.7 |
| CHIKV\|BA\|FS0055\|2015-07 | NB40 | FS0055 | 25.00 | BA | Feira de Santana | 7-2015 | M | 18706.0 | 87.9 | 941.5 | 96.9 | 98.4 |
| CHIKV\|BA\|FS0066\|2015-07 | NB41 | FS0066 | 22.00 | BA | Feira de Santana | 7-2015 | F | 16922.0 | 92.9 | 983.1 | 96.9 | 98.4 |
| CHIKV\|BA\|FS0089\|2015-07 | NB42 | FS0089 | 30.00 | BA | Feira de Santana | 7-2015 | F | 13283.0 | 69.0 | 812.2 | 96.9 | 98.1 |
| CHIKV\|BA\|FS0106\|2015-07 | NB43 | FS0106 | 30.00 | BA | Feira de Santana | 7-2015 | F | 15356.0 | 65.3 | 988.5 | 96.8 | 97.9 |
| CHIKV\|BA\|FS0116\|2015-07 | NB44 | FS0116 | 30.00 | BA | Feira de Santana | 7-2015 | F | 3426.0 | 31.8 | 455.9 | 96.8 | 97.7 |
| CHIKV\|BA\|FS0124\|2015-07 | NB45 | FS0124 | 30.00 | BA | Feira de Santana | 7-2015 | F | 14783.0 | 69.6 | 974.9 | 96.6 | 97.6 |
| CHIKV\|BA\|FS0129\|2015-07 | NB46 | FS0129 | 26.00 | BA | Feira de Santana | 7-2015 | M | 17350.0 | 76.1 | 1095.5 | 97.0 | 98.3 |
| CHIKV\|BA\|FS0132\|2015-07 | NB47 | FS0132 | 30.00 | BA | Feira de Santana | 7-2015 | F | 5605.0 | 50.1 | 482.5 | 96.4 | 97.3 |
| CHIKV\|BA\|FS0014A\|2015-08 | NB48 | FS0014A | 30.00 | BA | Feira de Santana | 8-2015 | F | 10910.0 | 62.1 | 763.4 | 96.3 | 97.3 |
| CHIKV\|BA\|DFS1686\|2019-10 | NB04 | DFS- 1686 | 22.57 | BA | Feira de Santana | 10-2019 | M | 314126.0 | 89.3 | 16143.3 | 96.6 | 97.9 |
| CHIKV\|BA\|DFS1786\|2019-11 | NB05 | DFS- 1786 | 30.60 | BA | Feira de Santana | 11-2019 | M | 219074.0 | 75.8 | 12052.6 | 96.4 | 97.5 |
| CHIKV\|BA\|DFS1808\|2019-11 | NB06 | DFS- 1808 | 28.60 | BA | Feira de Santana | 11-2019 | M | 326059.0 | 87.6 | 17001.9 | 96.5 | 97.9 |
| CHIKV\|BA\|DFS1837\|2019-11 | NB07 | DFS- 1837 | 21.72 | BA | Feira de Santana | 11-2019 | F | 207689.0 | 92.0 | 9717.2 | 96.7 | 98.1 |
| CHIKV\|BA\|DFS1849\|2019-11 | NB08 | DFS- 1849 | 29.54 | BA | Feira de Santana | 11-2019 | F | 253914.0 | 87.1 | 12450.4 | 96.6 | 98.0 |
| CHIKV\|BA\|DFS1854\|2019-11 | NB09 | DFS- 1854 | 35.25 | BA | Feira de Santana | 11-2019 | F | 262770.0 | 75.5 | 14107.5 | 96.6 | 97.6 |

| | | | | | | | | | | | | |
|---|---|---|---|---|---|---|---|---|---|---|---|---|
| CHIKV\|BA\|DFS1856\|2019-11 | NB10 | DFS- 1856 | 31.54 | BA | Feira de Santana | 11-2019 | F | 207524.0 | 82.7 | 10561.8 | 96.6 | 98.1 |

NT: Nucleotide
AA: Amino Aci

**Table S2. Centrality Metrics employed to measure transmission networks generated by the StrainHub tool.**

| Metastates | Degree Centrality | Indegree Centrality | Outdegree Centrality | Betweenness Centrality | Closeness Centrality | Source Hub Ratio |
|---|---|---|---|---|---|---|
| Paraguay | 1 | 1 | 0 | 0 | 0.07 | 0.00 |
| Southeast | 5 | 2 | 3 | 6 | 0.11 | 0.60 |
| Northeast | 6 | 2 | 4 | 9 | 0.13 | 0.67 |
| Haiti | 1 | 1 | 0 | 0 | 0.08 | 0.00 |
| Midwest | 4 | 2 | 2 | 3 | 0.11 | 0.50 |
| North | 2 | 1 | 1 | 0 | 0.08 | 0.50 |
| South | 1 | 1 | 0 | 0 | 0.07 | 0.00 |

**Table S3. Detailed site-by-site results from the BUSTED analysis for the envelope gene of clade I and II.**

| Codon | ER (ω>1)[1] | LogL | Epost[α][2] | LR[3] |
|---|---|---|---|---|
| **Clade I** | | | | |
| 187 | 187,712.29 | -78.412 | 0.294 | 12.721 |
| 372 | 742.301 | -31.012 | 1.07 | 6.651 |
| 699 | 344.611 | -92.661 | 0.294 | 3.369 |
| 60 | 211.964 | -79.268 | 0.294 | 2.566 |
| 62 | 57.168 | -17.817 | 3.408 | 4.991 |
| **Clade II** | | | | |
| 187 | 213,975.17 | -78.617 | 36.082 | 10.86 |
| 186 | 92,991.85 | -62.607 | 36.082 | 7.669 |
| 371 | 726.104 | -17.444 | 2.287 | 6.078 |
| 121 | 499.333 | -15.376 | 0.6 | 7.502 |

| | | | | |
|---|---|---|---|---|
| 184 | 105.634 | -59.121 | 35.724 | 0.507 |
| 29 | 96.931 | -36.048 | 33.526 | 4.941 |
| 60 | 19.358 | -78.757 | 36.082 | 1.279 |
| 370 | 12.692 | -42.632 | 36.074 | 1.966 |

[1]Evidence ratio for positive selection.
[2]Posterior mean of the synonymous rate, α.
[3]Site log-likelihood ratio contribution.

**Table S4. Sites under positive diversifying selection identified by HYPHY-MEME in the envelope gene of the clades I and II.**

| Site | alpha | beta+ | LRT | p-value |
|---|---|---|---|---|
| **Clade I** | | | | |
| 33 | 0 | 5.58 | 4.31 | 0.05 |
| 60 | 3.62 | 364.35 | 7.88 | 0.01 |
| 62 | 0 | 284.76 | 8.45 | 0.01 |
| 138 | 0 | 5.28 | 4.11 | 0.06 |
| 184 | 0 | 58.06 | 7.23 | 0.01 |
| 187 | 1.09 | 1521.76 | 21.03 | 0 |
| 282 | 0 | 2.31 | 3.43 | 0.09 |
| 372 | 0 | 6084.51 | 18.38 | 0 |
| 441 | 0 | 5.35 | 6.41 | 0.02 |
| 539 | 0 | 9.23 | 4.23 | 0.06 |
| 695 | 0 | 556.34 | 9.8 | 0 |
| 699 | 0 | 131.88 | 11.93 | 0 |
| **Clade II** | | | | |
| 29 | 0 | 315.97 | 9.55 | 0 |
| 53 | 0 | 3.7 | 3.71 | 0.07 |
| 60 | 4.18 | 469.33 | 4.83 | 0.04 |
| 121 | 0 | 181.66 | 8.77 | 0.01 |

| | | | | |
|---|---|---|---|---|
| 138 | 0 | 4.53 | 3.31 | 0.09 |
| 184 | 0 | 133.16 | 8.43 | 0.01 |
| 186 | 4.7 | 988.28 | 12.58 | 0 |
| 187 | 2.1 | 1124.28 | 20.44 | 0 |
| 371 | 0 | 256.38 | 9.31 | 0 |
| 432 | 0 | 4.87 | 3.88 | 0.07 |
| 441 | 0 | 3.16 | 3.38 | 0.09 |
| 539 | 0 | 70.36 | 4.82 | 0.04 |
| 699 | 0 | 9.05 | 3.67 | 0.08 |

alpha: synonymous substitution rate at a site.

beta+: non-synonymous substitution rate at a site for the positive selection category.

LRT: likelihood ratio test statistic for episodic diversification.
